# Assessing MRI Biomarker Repeatability to Guide Individualized TMS Treatment in Psychiatry

**DOI:** 10.64898/2026.07.25.26358908

**Authors:** Hosna Tavakoli, Reza Rostami, Alireza Fallahi, Narges Tabatabaei, Mohammad-Reza Nazem-Zadeh

## Abstract

**Background:** MRI has increasingly been explored as a biomarker for detecting structural and functional brain changes. For clinical decision-making, it is crucial to validate observed changes in MRI indices at the individual level. The uncertainty in longitudinal MRI indices can be quantified using the repeatability coefficient (RC).

**Methods:** Twenty healthy controls (10 males, 10 females) underwent two test-retest sessions of structural magnetic resonance imaging (MRI) and resting-state functional MRI (rs-fMRI) on the same day, separated by a 30-minute interval. RC values and their 95% confidence intervals (CI) were estimated for subcortical volumes, cortical thickness, and within-network functional connectivity. Additionally, 33 patients with mental health disorders underwent MRI before and after 20 sessions of transcranial magnetic stimulation (TMS). Percentage changes in MRI-derived indices were assessed at the individual level, with changes exceeding the RC threshold considered indicative of true change beyond measurement uncertainty.

**Results:** The RC showed measurement variability in subcortical volumetric in the range of 8% to 17.5% for caudate and left amygdala, respectively. For cortical thickness, the RC was measured between 3.5% and 16.5% for the left occipital pole and the left temporal pole, respectively. The RC% for fractional anisotropy (FA) measures were variable between 11.3% (the left middle cingulum) and 62.9% (the right anterior cingulum). For within-network connectivity, the RC was measured in a range of 9.7% and 29.4% for sensorimotor and visual networks, respectively. TMS-treated patients exhibited no changes beyond the RC in almost all subcortical volumes and within-network connectivity. The most frequent changes beyond the uncertainty were observed in FA measures, particularly in the posterior cingulum, where 17 out of 23 patients exhibited clinically meaningful alterations.

**Conclusion:** Structural brain features extracted from MRI demonstrated high reliability. Among all measures, FA, reflecting white matter integrity, was most sensitive in detecting neural changes following TMS, highlighting its potential utility as a treatment-responsive biomarker.

## 1 Introduction

Magnetic resonance imaging (MRI) serves as a valuable tool for measuring structural and functional alterations in the human brain for decades. Beyond that, MRI is increasingly regarded as a quantitative biomarker in neuroimaging research. Quantitative MRI metrics such as volumetric measurements, diffusion parameters including fractional anisotropy (FA), and functional network connectivity indices have been employed as biomarkers to assess disease progression, monitor treatment response, and facilitate disease classification in neurodegenerative and psychiatric disorders. MRI facilitates the longitudinal assessment of inherent anatomical and functional alterations associated with the aging process (Nair et al., 2026; Tang et al., 2023) as well as progression in neurodegenerative diseases such as Alzheimer’s disease (Li, Coyle, Maguire, Watson, & McGinnity, 2011; Lin et al., 2025; Matsuda, 2016), multiple sclerosis (Cagol et al., 2022), and Parkinson’s disease (Fereshtehnejad, Zeighami, Dagher, & Postuma, 2017; Scamarcia et al., 2022). In addition, MRI is frequently used to evaluate the effects of brain stimulation therapies, such as transcranial magnetic stimulation (TMS) and transcranial electrical stimulation (tES), and other neuromodulatory interventions on brain structure and function (Moraga-Amaro et al., 2023; Shen & Ward, 2021; T. Wang et al., 2024).

Although MRI is increasingly recognized as a promising biomarker, concerns remain regarding the validity of observed changes in MRI-derived measures, particularly at the individual level (Bashyam et al., 2020; Marek et al., 2022). This raises a fundamental question: what magnitude of change in an MRI measure constitutes a true or meaningful alteration? To address this, observed changes must exceed an expected level of uncertainty arising from factors such as image acquisition, processing, or natural biological and physiological variability. Only then can such changes be interpreted as reliable and clinically meaningful.

Uncertainty in the MRI-derived measures has been investigated in various studies using different metrics. Previous studies have addressed the concept of uncertainty and its potential applications in MRI analysis. Among MRI metrics, gray matter and white matter volumes are considered relatively reliable (Aryal, Chenevert, & Cao, 2016; Wüthrich et al., 2023). Uncertainty analysis in longitudinal imaging has been used to assess the validity of measurements for the assessment of radiation-induced neurotoxicity in patients with low-grade or benign tumors undergoing partial brain radiation therapy (Nazem-Zadeh, Chapman, Lawrence, Tsien, & Cao, 2013). Another notable application is the estimation of the uncertainty of interhemispheric change to identify the seizure focus in patients with temporal lobe epilepsy in a cross-sectional study (Nazem-Zadeh et al., 2014). With recent advancements in MRI acquisition, the reliability across different protocols is being investigated (Bardwell Speltz et al., 2025; Cahart et al., 2023).

In longitudinal neuroimaging studies, the repeatability coefficient (RC) quantifies the range of measurement uncertainty inherent to the imaging technique. While several studies have estimated RC for various diffusion MRI indices, reported values exhibit considerable variability (Schmida et al., 2026; Zou et al., 2025). This variability underscores that RC is influenced by multiple factors, including imaging acquisition parameters, data preprocessing pipelines, segmentation methodologies, and the intrinsic biological characteristics of the structures under investigation. Consequently, establishing a study-specific RC is a prerequisite for determining the reliability of observed longitudinal changes at the individual level. Such quantification is essential to assess whether measured changes exceed measurement noise, thereby informing their potential utility in clinical decision-making contexts.

Reproducibility, a core metric for assessing biomarker validity, quantifies the consistency of measurements when the same imaging protocol is repeated under stable conditions within a short time interval. In MRI research, reproducibility has been evaluated across numerous studies employing a range of statistical methods, including the intraclass correlation coefficient (ICC), coefficient of variation (CoV), and Bland–Altman limits of agreement (Botvinik-Nezer et al., 2020; Marek et al., 2022; Poldrack et al., 2017).

Test-retest MRI studies are designed to assess both the reliability and reproducibility of imaging-derived measures, thereby supporting the use of MRI in longitudinal research and clinical applications. It is shown that T1- and DTI-derived tissue metrics, including cortical and subcortical gray matter and white matter volume, exhibit insignificant mean differences both across different scanners and within-site repeat scans (Melzer et al., 2020). Other studies have assessed the reliability of task-based fMRI between scanners (Ibinson et al., 2022) or between sessions (Wüthrich et al., 2023) to further establish the robustness of the modality. Reproducibility has been evaluated using statistical metrics, such as the coefficient of variation (Bologna et al., 2023) or the correlation coefficient between test and retest measurements (Hu et al., 2023).

In this study, we estimated the RC for three distinct classes of MRI-derived measures: subcortical volumes, regional cortical thickness, and within-network functional connectivity. Using a test-retest dataset, we aimed to determine which measure exhibits the highest reliability for detecting longitudinal change. To quantify test-retest reproducibility, we computed the Pearson correlation coefficient (r) between corresponding measurements from the two sessions. Furthermore, to evaluate the potential clinical applicability of these reliability estimates, we applied the established RC thresholds to a separate cohort of 24 patients who underwent 20 sessions of TMS treatment. For each patient and measure, we compared the observed individual changes across the treatment period against the corresponding RC threshold to distinguish potential treatment-related changes from measurement variability.

## 2 Materials and Methods

### 2.1 Subjects

Twenty healthy volunteers and thirty-three patients with mental health disorders were recruited for this study. Exclusion criteria for all participants included a history of brain trauma, claustrophobia, and comorbid neurological conditions. Additionally, healthy controls were excluded if they had any current or past psychiatric disorders, while patients were excluded if they had comorbid psychiatric or neurological conditions. These criteria were reviewed, and the entire process was explained to participants over the phone.

After signing the consent form, healthy participants underwent two MRI sessions (test and retest), including structural and resting-state imaging on a single day at 30-minute intervals. This experimental design allowed us to assess the test-retest reliability and reproducibility of MRI-derived measures. All patients underwent TMS according to a protocol suggested by psychologists. MRI data acquired before treatment were analyzed to guide individualized TMS targeting. Each patient underwent a pre-treatment MRI session to assess structural and functional brain characteristics, using the same test-retest protocol applied to healthy controls. By comparing individual MRI profiles with those of healthy participants, personalized TMS protocols were developed to target the most affected brain regions. Patients were then assigned to one of five treatment groups based on these assessments: High-frequency left DLPFC, Low-frequency right DLPFC, High-frequency SMA, Low-frequency temporoparietal junction, or High-frequency left DLPFC/Low-frequency right DLPFC. Patients received 20 sessions of either high-frequency or low-frequency TMS over the course of 5 weeks. At the end of treatment, a follow-up MRI scan was conducted to evaluate TMS-induced neural changes. Depression and anxiety levels were assessed both before and after the intervention using the Beck Depression Inventory (BDI) and Beck Anxiety Inventory (BAI), respectively. These standardized self-report instruments provided a comprehensive evaluation of mood and anxiety symptom changes due to TMS.

### 2.2 MRI acquisition

Imaging was conducted on a 3T MRI system equipped with a 64-channel head coil (Prisma, Siemens, Erlangen, Germany) at the National Brain Mapping Laboratory, Tehran University, Iran, under the supervision of a neurologist (N.T.) present during all scans. Each session included a T1-weighted structural scan with the following protocol: TR = 1.9 s, TE = 2.26 ms, field of view (FOV) = 250 mm, matrix =256 × 256, sagittal plane, slice thickness=1 mm, voxel size= 1 x 1 x 1 mm, total of 176 slices. The resting-state scan lasted 396 seconds, acquired with the following parameters: TR = 1.2 s, TE = 30 ms, FOV=192 mm, matrix = 64 × 64, sagittal plane, slice thickness=3 mm, voxel size =3 x 3 x 3 mm, 42 slices.

### 2.3 MRI preprocessing

To extract structural measures, T1-weighted images were processed using FreeSurfer (v6.0.0; http://surfer.nmr.mgh.harvard.edu/). The processing steps included motion correction, intensity normalization, Talairach transformation, skull stripping, and anatomical segmentation. The cortical surface was parcellated into 78 regions (34 per hemisphere plus bilateral medial wall), and subcortical structures were segmented into 45 labeled regions. The most relevant cortical and subcortical regions from both hemispheres were selected for further analysis. Structural MRI measures included subcortical volumes of the thalamus, caudate, putamen, pallidum, hippocampus, and amygdala, and the cortical thickness of the fusiform gyrus, inferior parietal lobule (IPL), occipital cortex, frontal pole, temporal pole, and insula. These regions were selected based on their relevance in previous MRI studies. Quality control was conducted by visually inspecting cortical and subcortical segmentations across all healthy and patient datasets. No outliers were identified during this process.

Preprocessing of the rs-fMRI data was performed using the Data Processing and Analysis for Brain Imaging (DPABI) toolbox, an extension of the Data Processing Assistant for Resting-State fMRI (DPARSF) (Yan, Wang, Zuo, & Zang, 2016). The first 10 volumes were discarded, followed by slice-timing correction, motion realignment, brain extraction, and co-registration of the functional images to corresponding T1-weighted images. Subsequently, time series were extracted from 116 regions of interest (ROI) defined by the Automated Anatomical Labeling (AAL)atlas for both test and retest sessions. This resulted in a 320 × 116 time-series matrix per subject per session (320 representing the number of time points after discarding the first 10 volumes).

Functional connectivity within brain networks was derived from rs-fMRI data by calculating the Pearson correlation coefficients between pairs of regional time series. The analysis focused on brain networks defined by the 7-network parcellation from Yeo et al. (2011), and six functional networks based on the AAL atlas (Tzourio-Mazoyer et al., 2002). The Yeo networks included the Default Mode Network (DMN), Visual Network (VSN), Dorsal Attention Network (DAN), Frontoparietal Network (FPN), Sensorimotor Network (SMN), Limbic Network (LIN), and Ventral Attention Network (VAN). The AAL networks comprised the Default Mode, Auditory (AUN), Visual, Attention (ATN), Sensorimotor, and Subcortical (SCN) networks. A comprehensive list of network definitions and their constituent regions is provided in Supplementary Table S1. Within-network connectivity was computed as the mean functional connectivity across all pairs of regions within each network."

### 2.4 Repeatability Coefficient

There are several motivations for quantifying the uncertainty of MRI-derived measures. In this study, we focus on two primary objectives: (1) to assess the validity and reliability of the MRI measures, and (2) to determine whether an observed change in a parameter reflects a true biological or clinical change beyond measurement variability.

To estimate the level of uncertainty, we calculated the RC values for each MRI-derived features as follows:

Let *Y*_*ik*_ denote the observed value of a specific MRI measure for the *i*^*th*^ subject and *k*^*th*^ replication, where k=1, 2 and n=20 in our test-retest dataset.

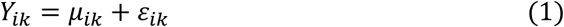

where *μ*_*i*_ is the true value for subject i, and *ε*_*ik*_ is the within-subject error. The within-subject variance is given by 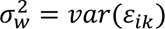 and the between-subject variance is 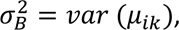 followinga normalized one-way ANOVA model. The total variance is 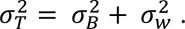 The within-subject mean square (WMS) and between-subject mean square (BMS) are estimated using one-way ANOVA model as:

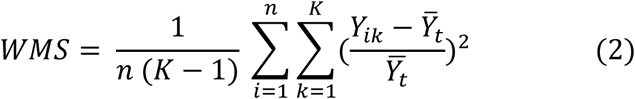

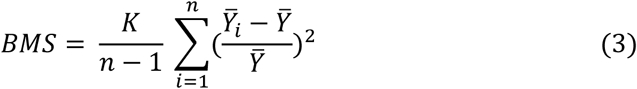

where <u>*Y*_*i*_</u> is the mean of replications for the *i*^*th*^ subject, and *Ȳ* is the overall mean across all subjects and sessions.

The within-subject variance is estimated as 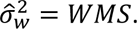 For the case where *K* = 2 (test and retest), Equation (2) can be rewritten as:

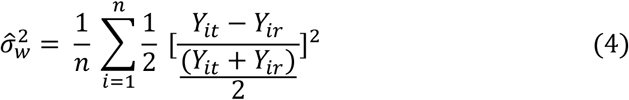

where t and r denote test and retest, respectively.

The RC is then defined as 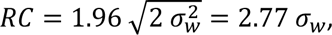 which means the difference between two measurements for the same subject is expected to lie within the interval -RC to RC for 95% of the subjects.

Assuming WMS follows a chi-squared distribution: 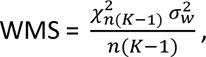 the 95% CI of the estimated RC for *K* = 2 is given by:

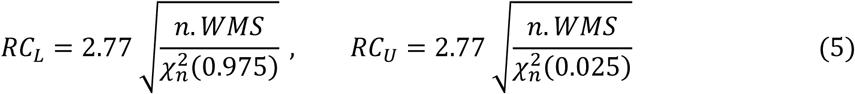

Thus, the estimated repeatability coefficient 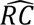 falls within its 95% CI:

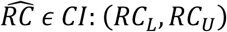

where 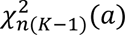 is the *a*^*th*^ percentile of the chi-squared distributions *n* (*K* − 1) degrees of freedom.

Assuming there is no true biological change in a structure between test and retest, such as that which might occur due to disease progression or treatment, any observed difference must be attributed to random and/or systematic errors. These errors may arise from variability introduced by the imaging device, acquisition parameters, patient repositioning, image processing and analysis steps, or subject-specific biological, physical, and physiological fluctuations (Barnhart & Barboriak, 2009).

The F-test is defined as the ratio of BMS to WMS and is used to assess whether the variance between subjects is significantly greater than the variance within subjects. A significant result would suggest that the within-subject variability is small compared to between-subject variability, supporting the stability and reliability of the measurement.

### 2.5 Level of longitudinal changes

In group analyses, the presence of a change is typically determined using statistical methods to identify generalizable effects across the population. However, while group-level results may demonstrate the effectiveness, or lack thereof, of a given intervention, individual-level variability, including opposite or inconsistent outcomes, can be obscured. To address this limitation, we assessed whether post-versus pretreatment changes in each MRI-derived measure, across brain structures and networks, represented true individual-level changes.

Patients underwent MRI sessions both before and after completing TMS treatment. To ensure the reliability of observed changes, we assessed whether each change exceeded the expected limits of measurement variability, defined by the RC. Specifically, a change was considered meaningful only if it fell outside the interval (-RC, RC). For a more conservative comparison, we consider the interval (-*RC*_*L*_, *RC*_*L*_) for as the ground to compare the changes with. Three outcome categories were defined:

1. No change: If the calculated change for a given measure fell within this interval, it was interpreted as not exceeding measurement uncertainty, and thus, no meaningful change was assumed.
2. Positive change: If the change (Δ*Y*) exceeded the upper bound of the interval (+*RC*_*L*_), it was classified as a positive response to treatment.
3. Negative change: Conversely, if the change fell below the minus of lower bound (-*RC*_*L*_), it was categorized as a negative response.

We then calculated the proportion of true changes observed for each brain feature and investigated whether these neural changes were associated with variations in patients’ depression and anxiety scores following treatment. To evaluate these associations, Pearson’s correlation coefficients and linear regression analyses were employed.

### 2.6 Statistical analysis

To assess statistical differences between test and retest measurements, paired-sample t-tests were performed for each brain region and network. To assess the reproducibility of MRI features, correlation coefficients were calculated between test and retest measurements for both structural and functional MRI metrics.

Due to the unequal distribution of females and males in the sample, with males comprising the majority of participants, a two-way repeated measures ANOVA was performed to assess pre- to post-treatment. To further investigate the relationship between symptom severity and neural changes, participants were stratified into subgroups based on their baseline BDI and BAI scores. For depression, individuals with BDI scores less than 20 were classified as moderate, while those with scores above 20 were categorized as severe. Similarly, for anxiety, participants with BAI scores below 15 were categorized as moderate, and those with scores above 15 as severe. Within each subgroup, we calculated the correlations between changes in MRI measures and: (1) baseline BDI and BAI scores, and (2) changes in BDI and BAI from pre- to post-treatment.

## 3 Results

### 3.1 Subjects

The demographic characteristics of the patient cohort are summarized in **Error! Reference source not found.**. Nine patients were excluded from the study at various stages of treatment, resulting in a final sample of 24 participants (16 males, 8 females). The mean age of the participants was 34.8 years (± 12.6), indicating a relatively wide age range. In the RC calculations for FA, one healthy subject was excluded as an outlier. Additionally, in the true change analysis, one patient was excluded due to missing resting-state and DTI data.

In terms of pre-treatment symptom severity, the mean BDI score was 23.9 (± 11.2) and the mean BAI score was 22 (± 14.6), reflecting moderate levels of both depression and anxiety before treatment. Given the unequal distribution of male and female participants, with males representing the majority of the sample, a two-way repeated measures ANOVA was performed with Time (pre- vs. post-treatment) as a within-subject factor and Gender (female, male) as a between-subject factor. As it is shown in Table 2, for depression scores, the analysis revealed a significant main effect of Time, F(1, 22) = 15.67, p = 0.0007, indicating a marked reduction in depressive symptoms following treatment. Neither the main effect of Gender, F(1, 22) = 0.80, p = 0.38, nor the Time × Gender interaction, F(1, 22) = 0.18, p = 0.68, reached significance, suggesting that both females and males experienced comparable decreases in depression. For anxiety scores, a significant main effect of Time was also observed, F(1, 22) = 17.38, p = 0.0004. Importantly, a significant Time × Gender interaction emerged, F(1, 22) = 4.58, p = 0.044, indicating that the magnitude of anxiety reduction differed between genders.

**Table 1.** Demographic and clinical characteristics of the patients. BDI, Beck Depression Inventory; BAI, Beck Anxiety Inventory.

| Variable | Mean ± SD | t-value | p-value |
| --- | --- | --- | --- |
| Gender (M/F) | 16 / 8 | — | — |
| Age (years) | 34.8 ± 12.6 | — | — |
| BDI-pre | 23.9 ± 11.2 | 4.016 | <0.001*** |
| BDI-post | 15.8 ± 9.8 | | |
| BAI-pre | 22 ± 14.6 | 3.596 | 0.001** |
| BAI-post | 13.5 ± 10.9 | | |

**Table 2.** Two-way repeated measures ANOVA results for depression (BDI) and anxiety scores. Time represents pre- vs. post-treatment measurements, and Gender represents females and males.

| Measure | Effect | F (df1, df2) | p-value |
| --- | --- | --- | --- |
| Depression | Time | 15.67 (1,22) | <0.001*** |
|  | Gender | 0.80 (1,22) | 0.38 |
| | Time × Gender | 0.18 (1,22) | 0.68 |
| Anxiety | Time | 17.38 (1,22) | <0.001*** |
|  | Gender | 1.37 (1,22) | 0.25 |
| | Time × Gender | 4.58 (1,22) | <0.05* |

After controlling for gender, the change in depression scores remained statistically significant (adjusted ΔBDI = −9.1, p = 0.009), indicating a treatment effect independent of gender. Despite a significant TIME × GENDER interaction, the overall change in anxiety scores remained significant after controlling for gender (adjusted ΔBAI = −14.0, p < 0.001), indicating gender-dependent magnitude but preserved treatment effect.

### 3.2 Reliability and reproducibility of MRI measures

We quantified the differences between test and retest data for five MRI-derived brain features: subcortical volume, cortical thickness, fractional anisotropy (FA), and within-network functional connectivity from both the AAL and Yeo atlases. Boxplots illustrating these comparisons are presented in Supplementary Figure S1.

In the initial analysis, no significant test-retest differences were observed in within-network connectivity based on either the AAL or Yeo network definitions. For cortical thickness, a statistically significant difference was found in the fusiform gyrus with p = 0.001 and p = 0.041 for the left and the right fusiform gyri, respectively In the FA metric, a significant difference was observed in the right anterior cingulate region (p = 0.017) between the test and retest sessions.

Pearson correlation coefficients between test-retest values for all MRI measure are presented in Figure 1. Structural features (subcortical volume, cortical thickness, FA) showed higher correlation values compared to functional measures (within-network connectivity), suggesting greater test-retest reliability for structural metrics.

**Figure 1.**
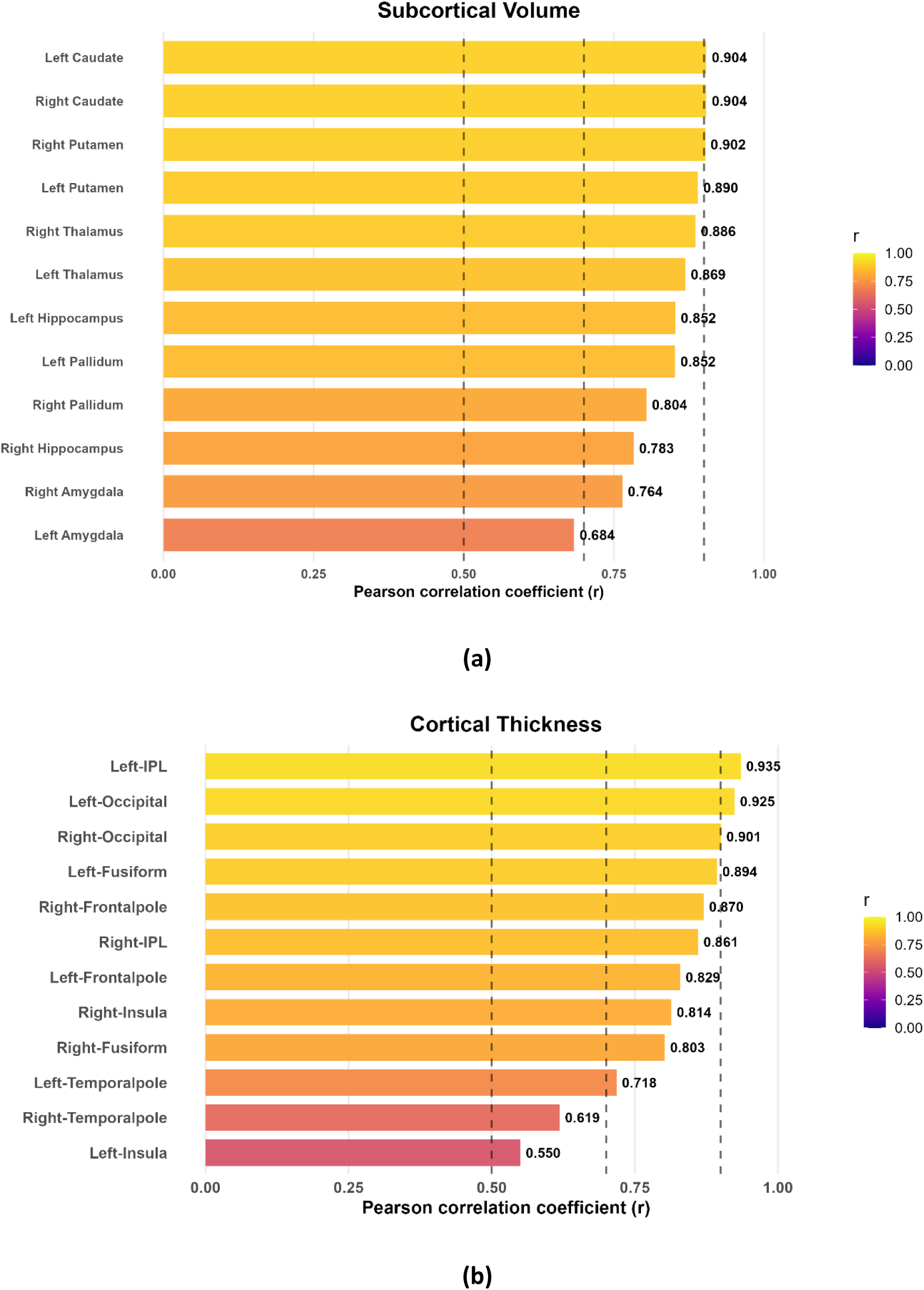
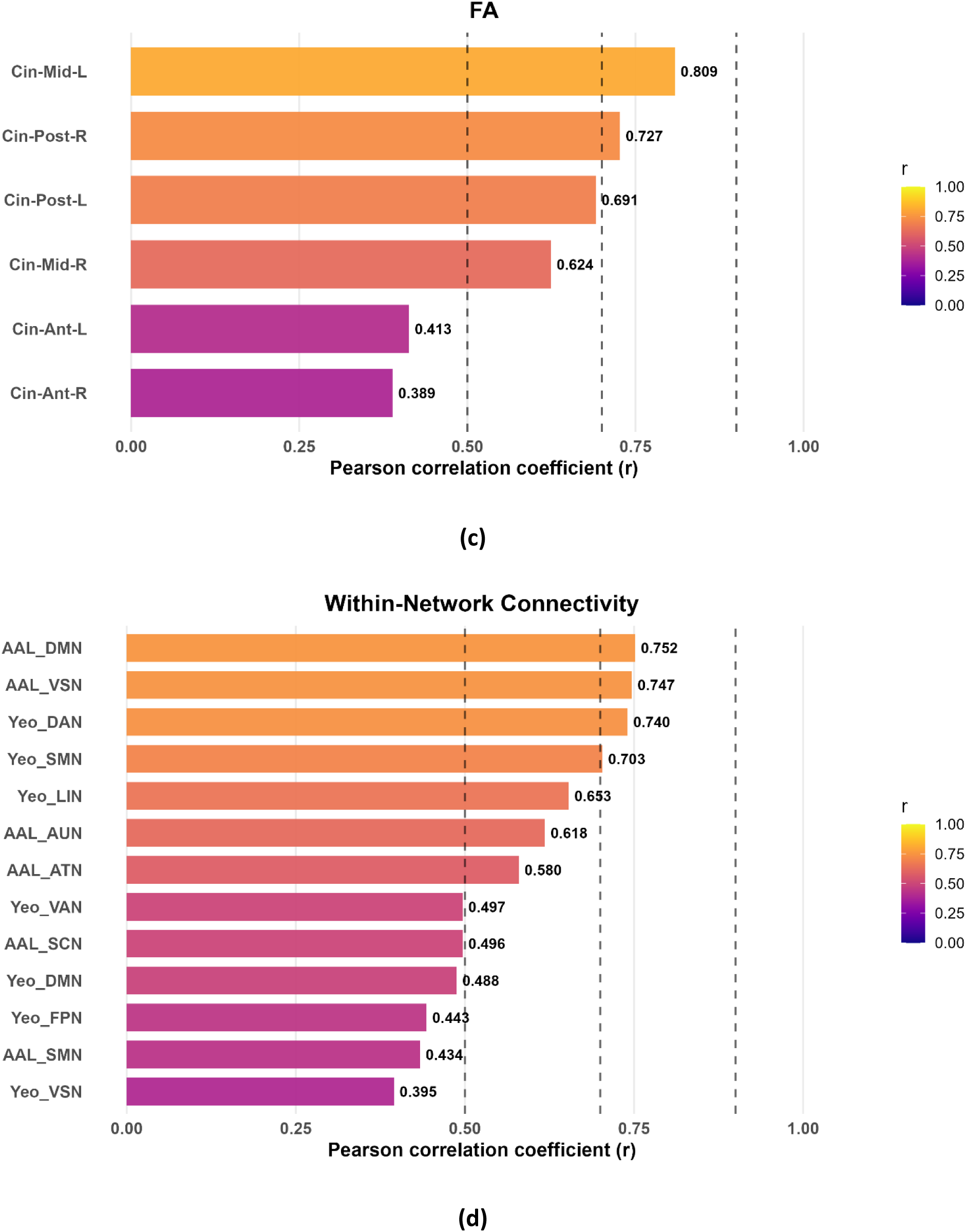
Pearson correlation coefficients for test–retest reproducibility of MRI-derived measures: (a) subcortical volume, (b) cortical thickness, (c) fractional anisotropy (FA), and (d) within-network functional connectivity in AAL and Yeo networks. These correlations represent the reproducibility of each MRI feature across sessions. L, left; R, right; IPL, inferior parietal lobule; DMN, default mode network; VSN, visual network; DAN, dorsal attention network; FPN, frontoparietal network; SMN, sensorimotor network; LIN, limbic network; AUN, auditory network; ATN, attention network; SCN, subcortical network.

Since the within-subject variance was considerably smaller than the between-subject variance across all MRI parameters, the dataset was deemed valid for estimation of RC. Using test-retest data from 20 healthy subjects, we quantified the uncertainty in three MRI-derived measures: subcortical volume, cortical thickness, and within-network functional connectivity. The estimated RC for structural and functional measures, along with the corresponding lower (*RC*_*L*_) and upper (*RC*_*U*_) confidence bounds, are summarized in Figure 2 and Table S2.

**Figure 2.**
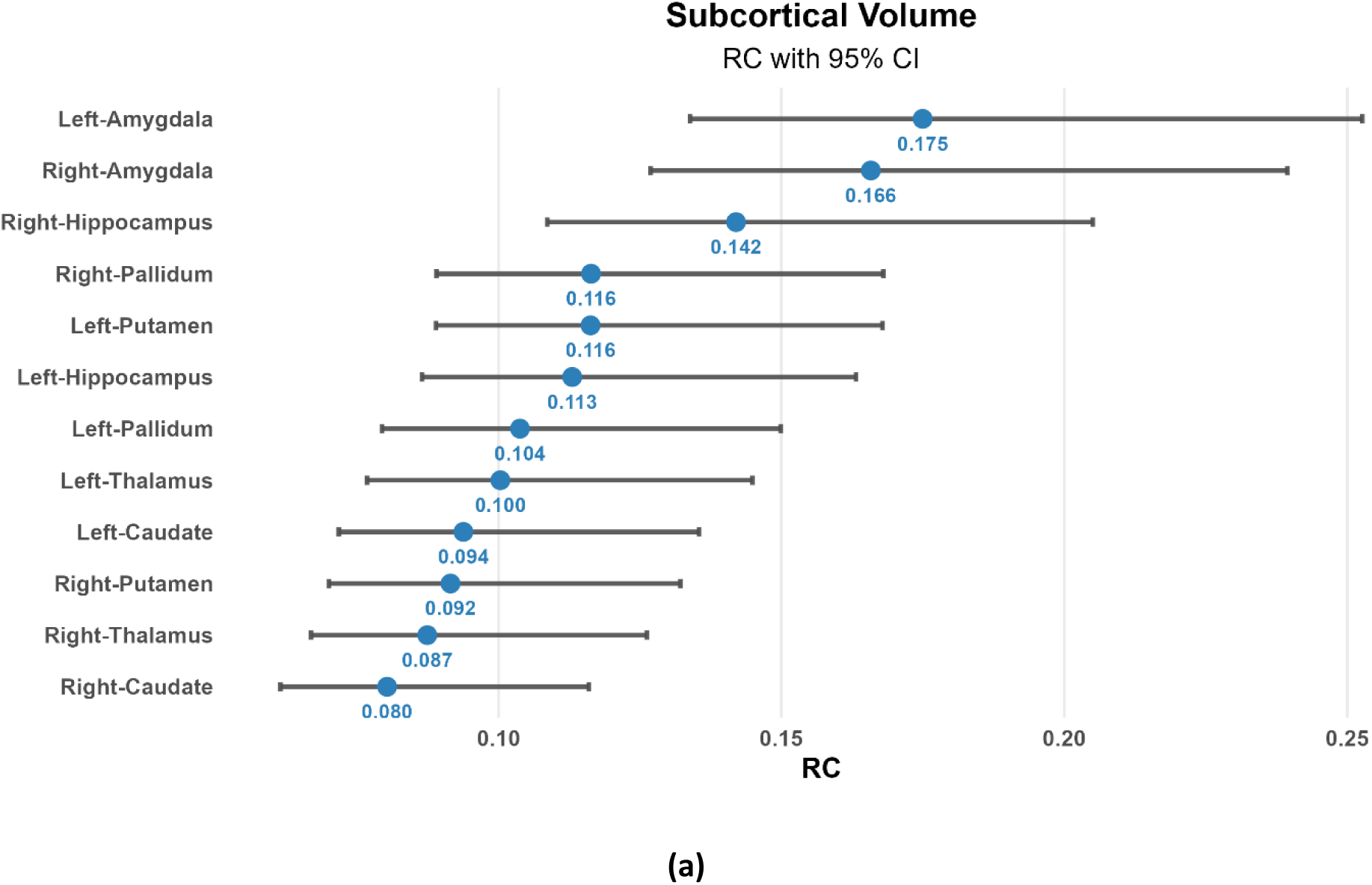
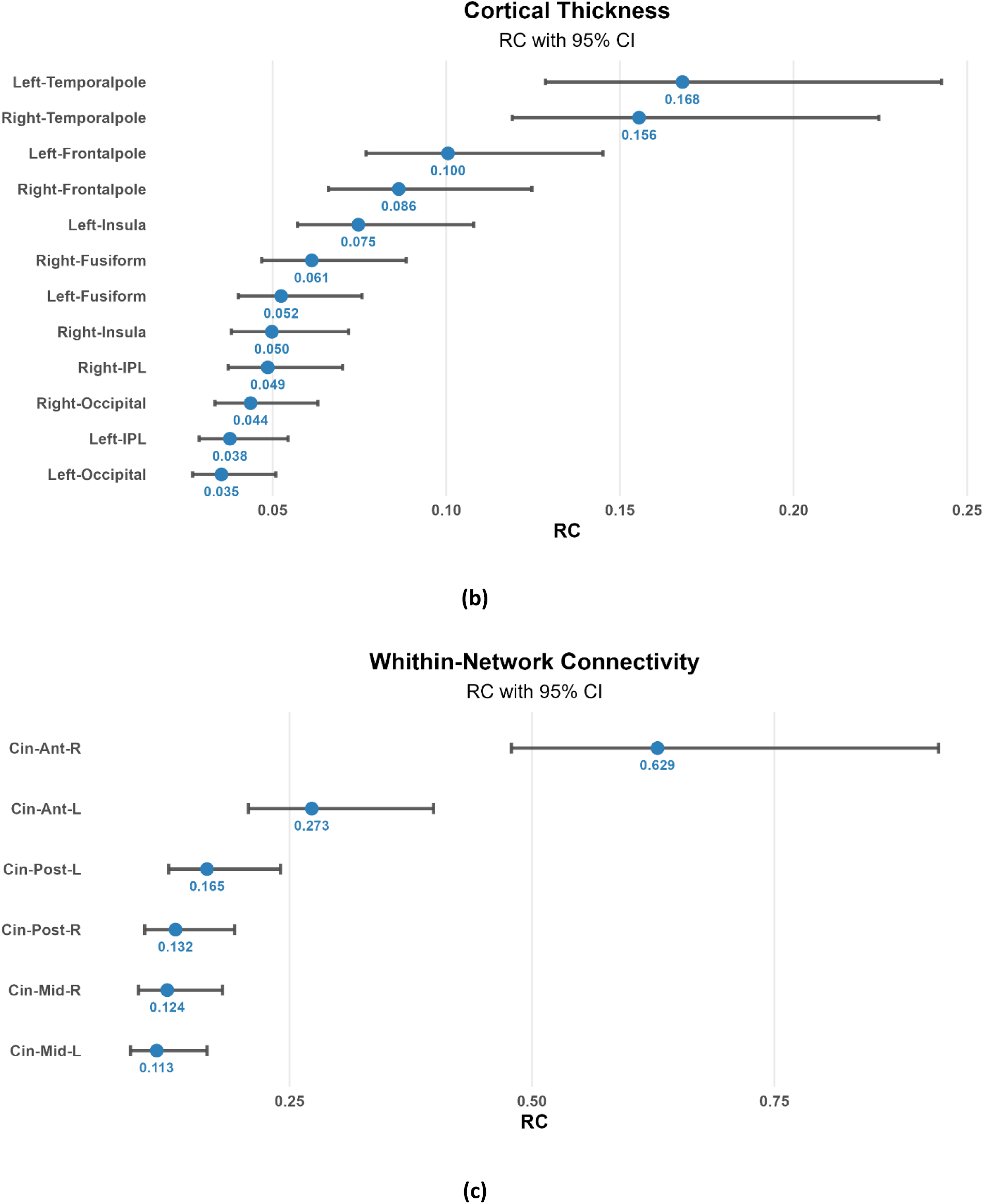
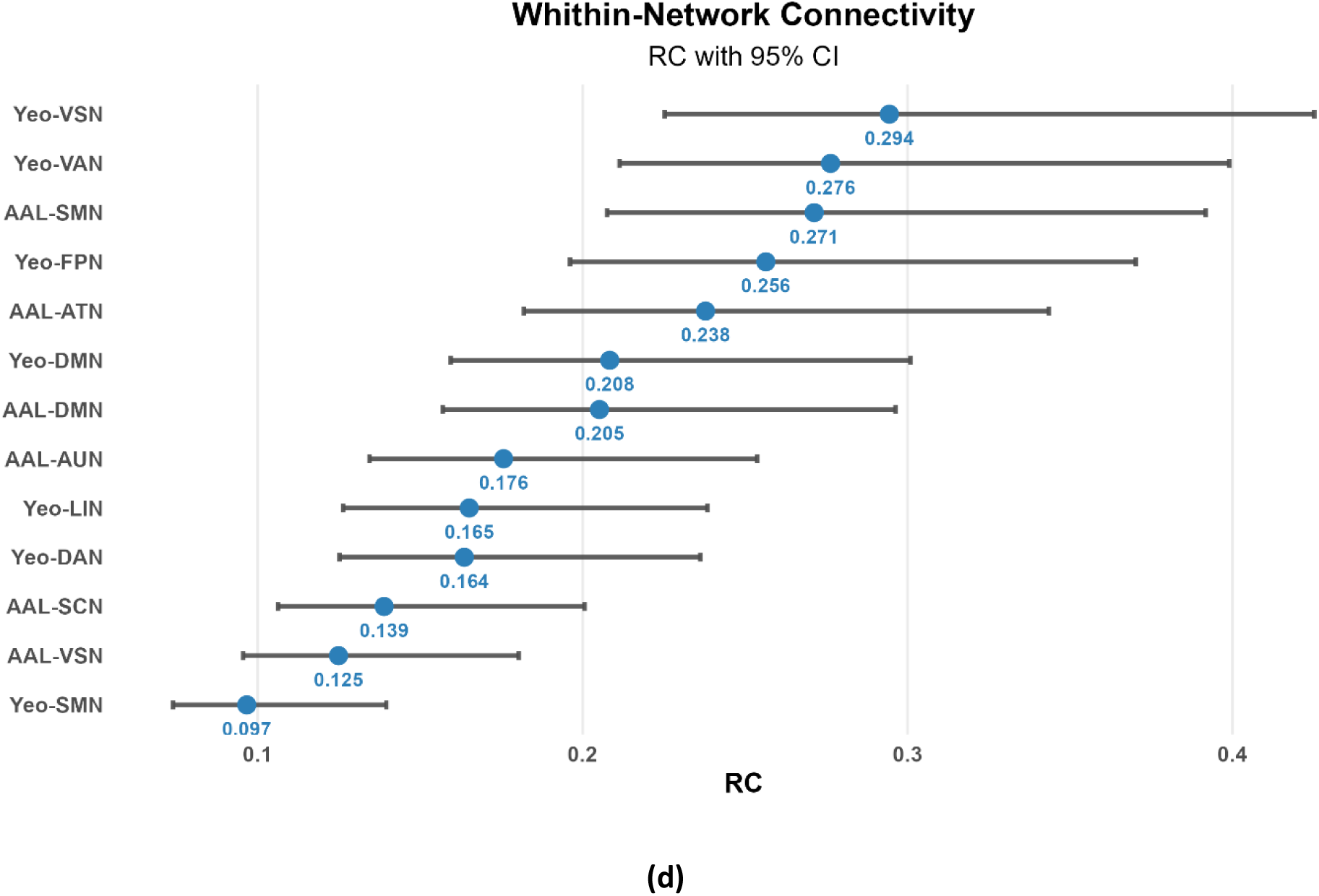
Repeatability Coefficients (RC) with (*RC*_*L*_, *RC*_*U*_) intervals are shown for each measure: (a) subcortical volume, (b) cortical thickness, (c) fractional anisotropy (FA), and within-network functional connectivity in AAL and Yeo networks (d). Higher RC values indicate larger expected differences between repeated measurements, while smaller RC values reflect higher reproducibility.

For subcortical volumes, the smallest RC% was observed in the right caudate (8%), while the largest RC% was found in the left amygdala (17.5%). For cortical thickness, the left occipital region exhibited the smallest RC% (3.5%), and the left temporal pole showed the largest (16.8%). The RC% values for FA values ranged from 11.3% in the left middle cingulum to 62.9% in the right anterior cingulum.

Within-network functional connectivity based on AAL networks had RC% values ranging from 12.5% in the visual network to 27.1% in the sensorimotor network. Similarly, in Yeo networks, RC% values ranged from 9.7% in the sensorimotor network to 29.4% in the visual network.

Figure 3 illustrates the lower and upper bounds of the RC (*RC*_*L*_ and *RC*_*U*_) along with the observed changes in MRI-derived measures from the test-retest dataset of 20 healthy subjects. Two subjects exhibited extreme deviations in their FA values and were excluded from the FA analysis as statistical outliers. For nearly all remaining subjects, the observed changes across the five MRI features, subcortical volume, cortical thickness, FA, and within-network connectivity based on AAL and Yeo networks, fell within the RC bounds. This consistency indicates high measurement reliability and supports the use of these features in detecting meaningful longitudinal changes.

**Figure 3.**
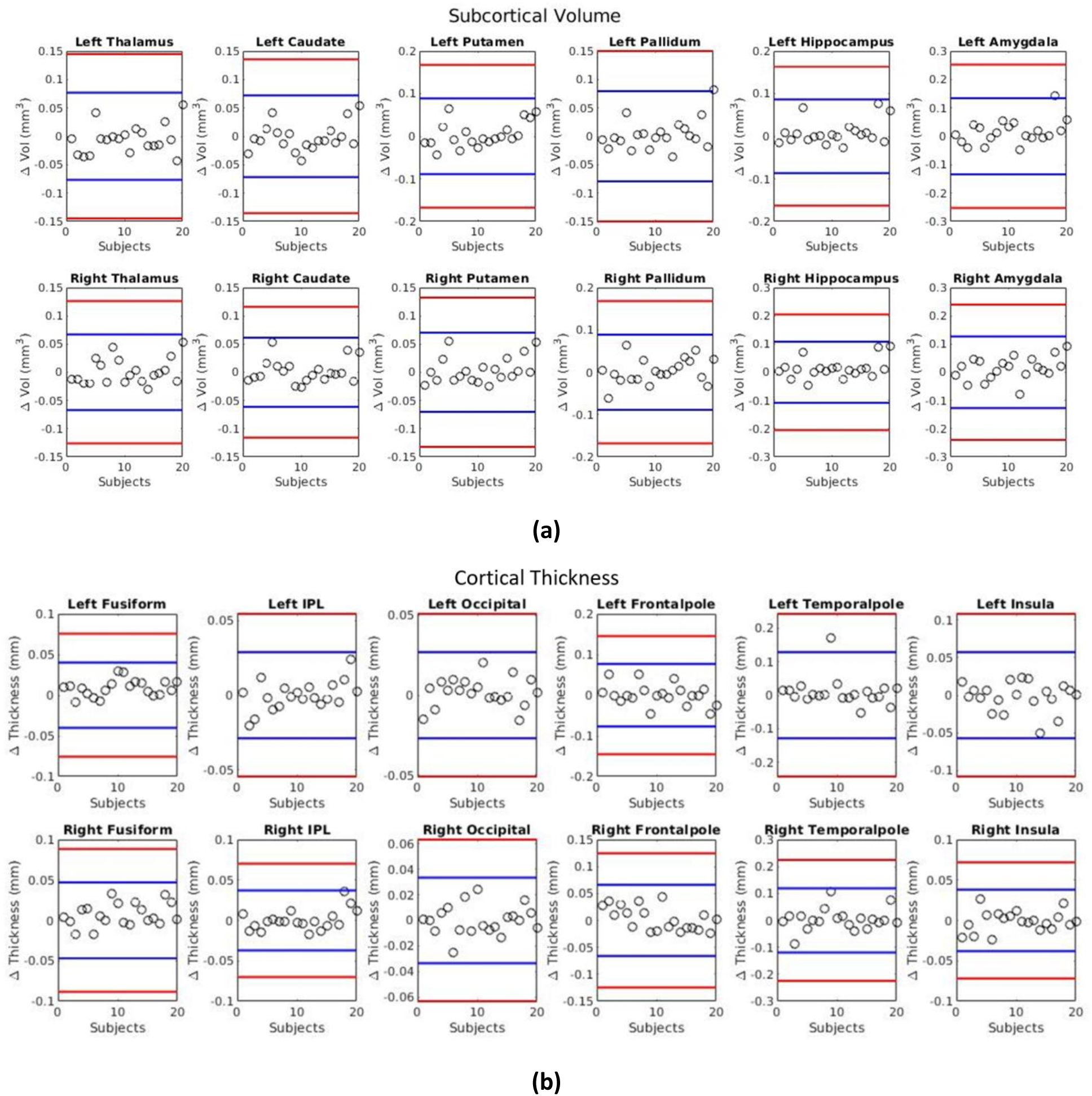
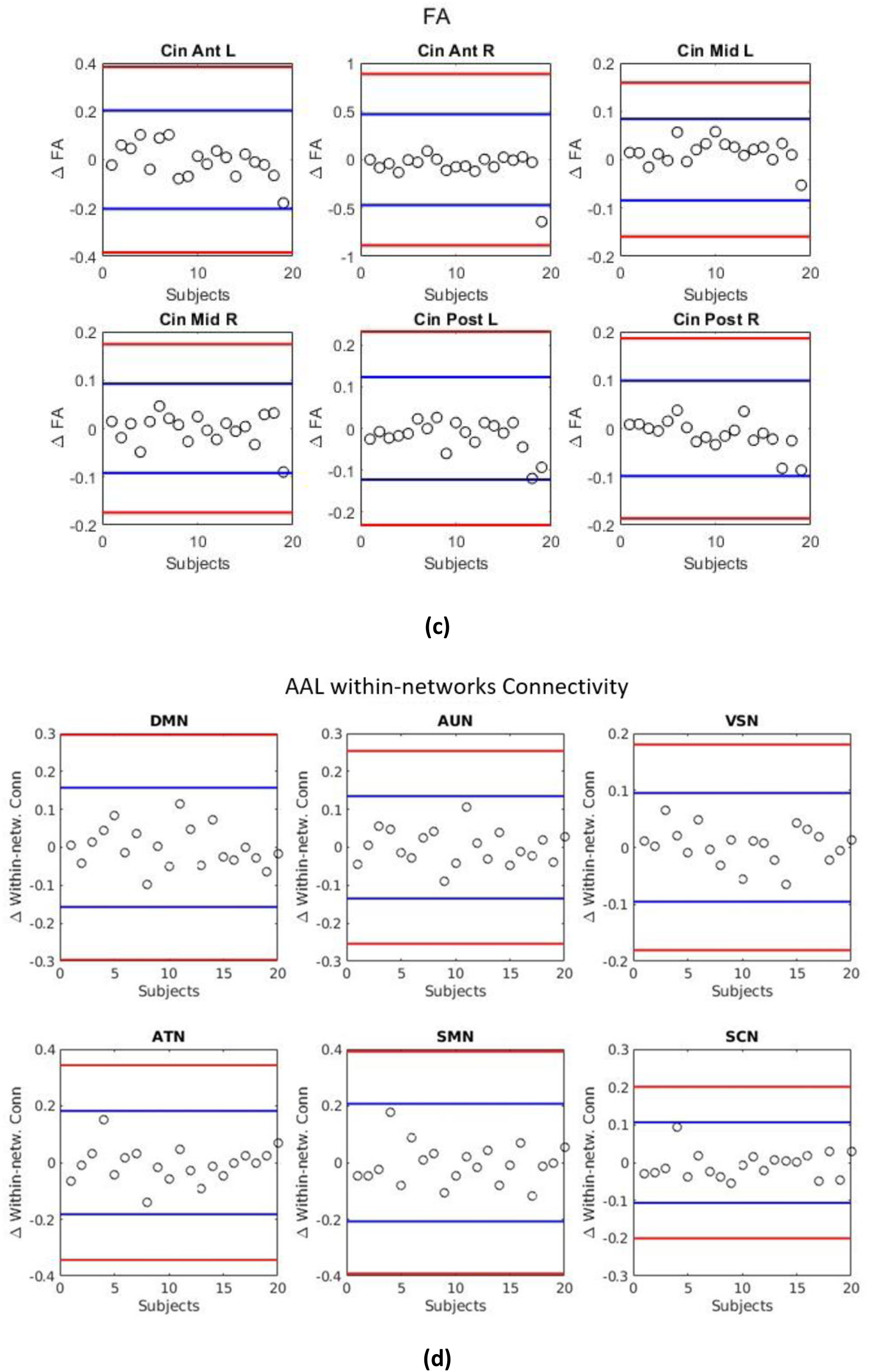
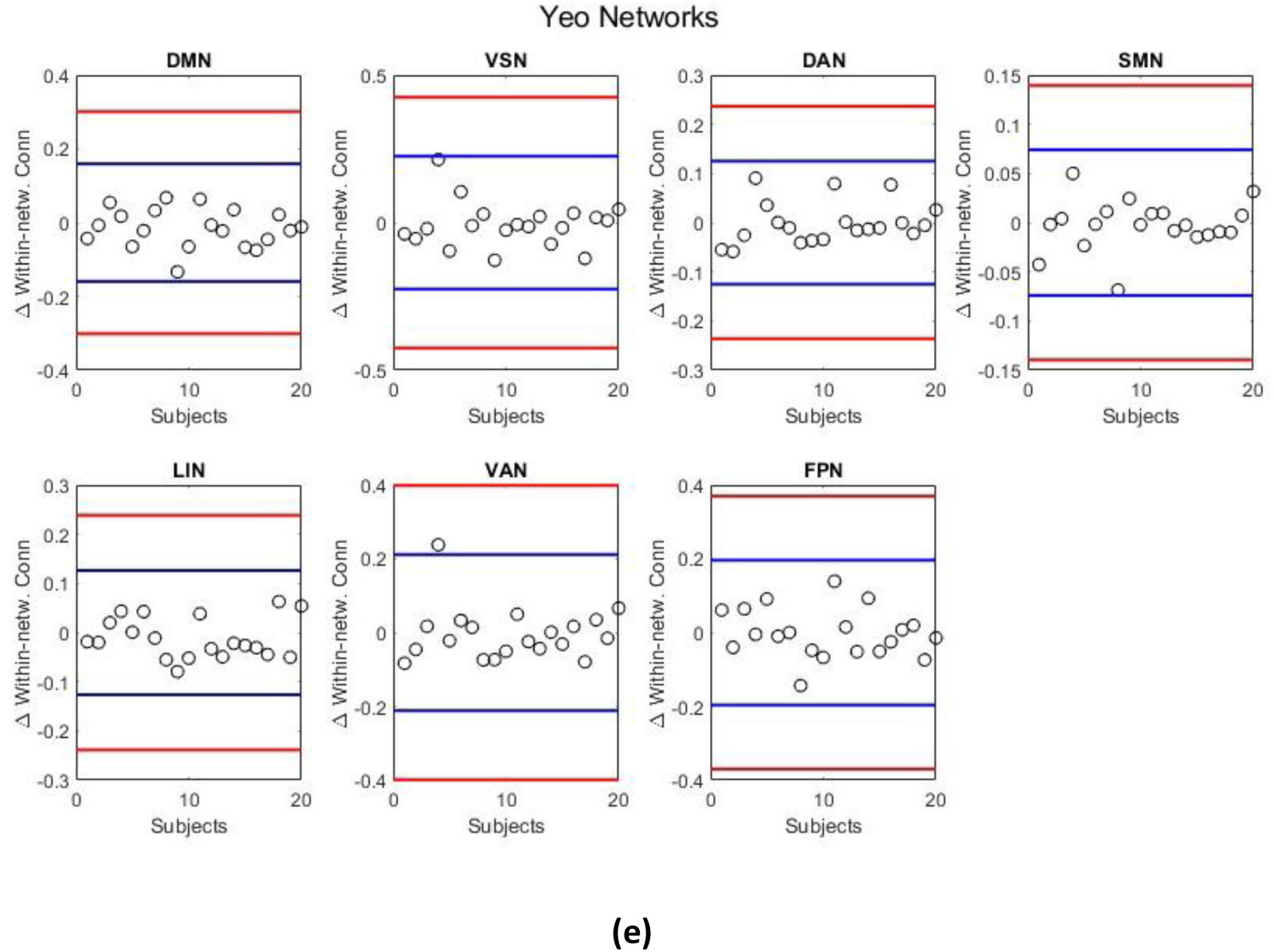
Lower and upper bounds of the RC with observed changes in test-retest data from healthy subjects (shown as circles) for: (a) subcortical volumes, (b) cortical thickness, (c) fractional anisotropy (FA), (d) within-network connectivity based on AAL networks, and (e) within-network connectivity based on Yeo networks. The blue lines indicate the inner RC interval (−*RC*_*L*_, *RC*_*L*_), and red lines represent the outer RC interval (−*RC*_*U*_, *RC*_*U*_). Abbreviations: IPL, inferior parietal lobule; DMN, default mode network; VSN, visual network; DAN, dorsal attention network; FPN, frontoparietal network; SMN, sensorimotor network; LIN, limbic network; AUN, auditory network; ATN, attention network; SCN, subcortical network.

### 3.3 Longitudinal changes of MRI measures among patients

Table 3 presents the percentage of patients whose FA values exceeded the uncertainty range defined by the interval (−*RC*_*L*_, *RC*_*L*_), as FA demonstrated a high number of true changes among all MRI-derived features. To ensure a conservative estimation, the lower bound of the RC interval was used in the classification of change.

**Table 3.** The number of true changes in FA values.

| White matter regions | Patients with true changes in FA,<br>n (%) |
| --- | --- |
| Left Anterior Cingulum (Cingulum-Ant-L) | 9 (39.1%) |
| Right Anterior Cingulum (Cingulum-Ant-R) | 2 (8.69%) |
| Left Middle Cingulum (Cingulum-Mid-L) | 9 (39.1%) |
| Right Middle Cingulum (Cingulum-Mid-R) | 8 (34.8%) |
| Left Posterior Cingulum (Cingulum-Post-L) | 17 (73.9%) |
| Right Posterior Cingulum (Cingulum-Post-R) | 17 (73.9%) |

**Note:** True changes are defined as FA values that exceed the uncertainty based on test-retest repeatability thresholds derived from healthy control data.

Patients with values falling outside the uncertainty range, either above or below (−*RC*_*L*_, *RC*_*L*_), were considered to exhibit true longitudinal changes. Figure 4 (a-d) illustrates the individual change scores (Δ*Y*_*t*_) for each patient across subcortical volumes, cortical thickness, FA, and within-network functional connectivity. The highest frequency of true changes was observed in the posterior cingulate gyrus, where 17 out of 23 patients exhibited changes beyond the defined uncertainty range.

**Figure 4.**
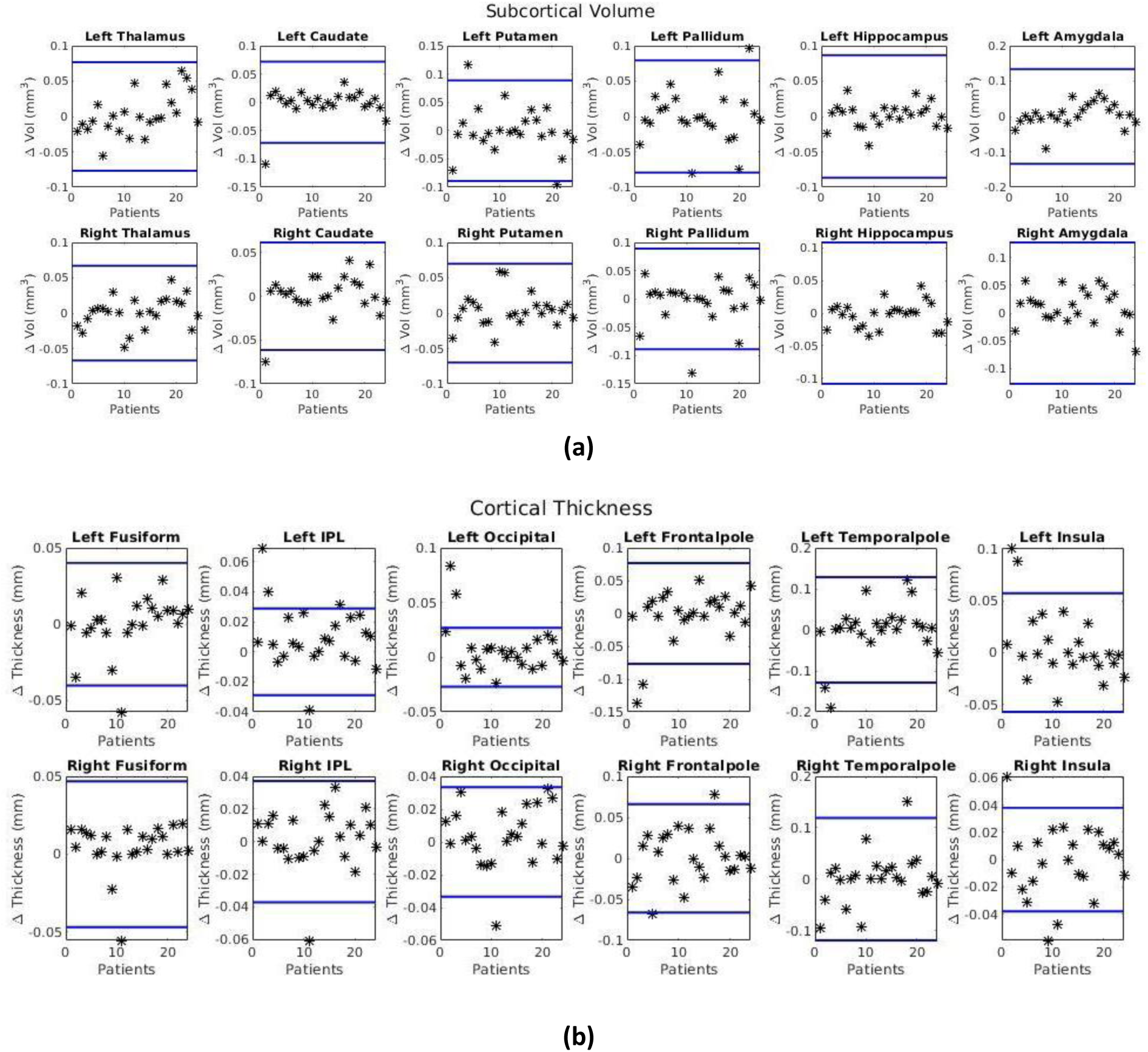
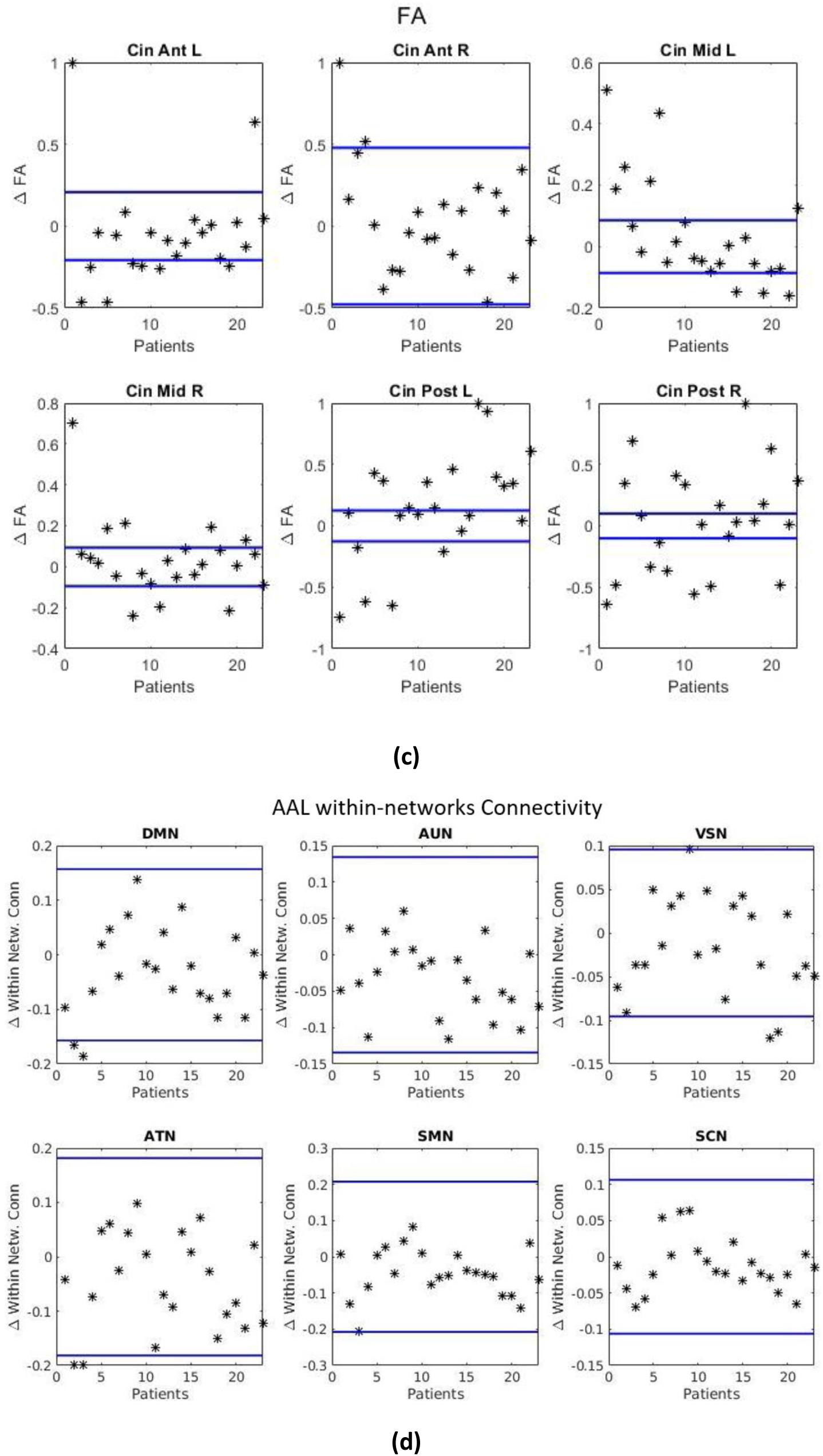
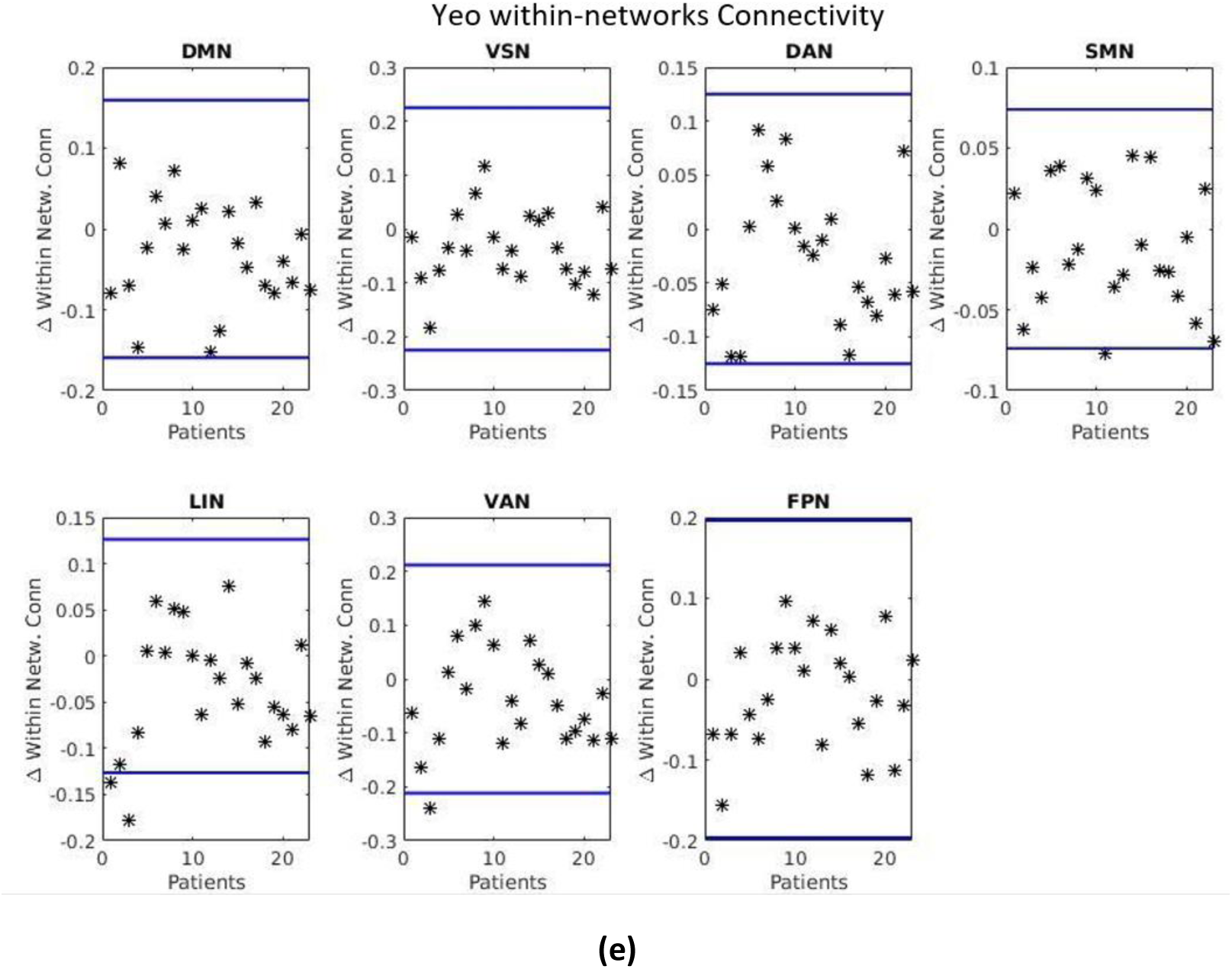
The individual-level changes in MRI-derived measures for all 24 patients (shown as stars), plotted against the lower repeatability interval (−RC*L*, RC*L*) (−RCL, RCL), are represented by blue lines. The panels show changes in: (a) subcortical volumes, (b) cortical thickness, (c) fractional anisotropy (FA), (d) within-network connectivity based on AAL networks, and (e) within-network connectivity based on Yeo networks. Most of the observed patient-level changes fall within the uncertainty bounds, while specific regions (e.g., the posterior cingulum in FA) show a higher frequency of changes beyond the lower RC threshold. Abbreviations: IPL, inferior parietal lobule; DMN, default mode network; VSN, visual network; DAN, dorsal attention network; FPN, frontoparietal network; SMN, sensorimotor network; LIN, limbic network; AUN, auditory network; ATN, attention network; SCN, subcortical network.

### 3.4 Correlation between changes in brain features and depression and anxiety

Table 4 presents the demographic and baseline characteristics of the study groups. No significant differences were observed in age or gender distribution between groups, whereas baseline BDI and BAI scores differed significantly between the moderate and severe depression and anxiety groups, respectively.

**Table 4.** Demographic data and baseline BDI and BAI scores of patients classified into moderate and severe subgroups.

| Grouping Criterion | Subgroup | n | Gender (F/M) | Age (years)<br>Mean ± SD | BDI-pre<br>Mean ± SD | BAI-pre<br>Mean ± SD | p-value†<br>(BDI) | p-value†<br>(BAI) | p-value†<br>(Age) |
| --- | --- | --- | --- | --- | --- | --- | --- | --- | --- |
| BDI Group | Moderate | 8 | 3 / 5 | 35.3 ± 10.5 | 17.3 ± 12.2 | 11.6 ± 6.0 | 0.000* | 0.300 | 0.90 |
| | Severe | 15 | 6 / 9 | 34.5 ± 14.3 | 24.1 ± 15.9 | 30.7 ± 7.1 | | | |

| Grouping<br>Criterion | Subgroup | n | Gender<br>(F/M) | Age (years)<br>Mean ± SD | BDI-pre<br>Mean ± SD | BAI-pre<br>Mean ± SD | p-value†<br>(BDI) | p-value†<br>(BAI) | p-value†<br>(Age) |
| --- | --- | --- | --- | --- | --- | --- | --- | --- | --- |
| BAI Group | Moderate | 8 | 1 / 7 | 34.8 ± 12.9 | 21.8 ± 12.9 | 6.4 ± 3.9 | 0.55 | 0.000* | 0.93 |
| | Severe | 15 | 8 / 7 | 35.3 ± 13.1 | 24.8 ± 10.9 | 30.8 ± 10.7 | | | |

The relationship between baseline and changes in depression and anxiety severity with subsequent changes in brain features was evaluated using Pearson’s correlation coefficient in each severe and moderate groups. Supplementary Figures S2 and S3 show scatter plots illustrating the associations of baseline depression and anxiety scores with changes in MRI-derived measures, and Figures S4 and S5 show the relationship between changes in depression and anxiety scores and corresponding changes in MRI measures, respectively. After applying a Bonferroni correction for multiple comparisons, two statistically significant correlations remained for the patients with severe anxiety: (1) a positive association between change in BAI scores and change in within-network connectivity of DMN in Yeo (r = 0.78, p = 0.001), (2) a positive correlation between pre-treatment BAI and the change in cortical thickness of right frontal pole (r = 0.68, p = 0.005). This suggests that patients with higher anxiety experienced greater alterations in brain structure and function following treatment.

## 4. Discussion

Reliable neuroimaging biomarkers are essential for guiding individualized psychiatric treatments, and their repeatability and reproducibility must be thoroughly evaluated before clinical application. Over the past decade, MRI has become a widely adopted modality for identifying both disease-related and treatment-induced brain changes in psychiatric and neurobiological disorders (Leocani, Rocca, & Comi, 2016; Tusconi & Dursun, 2025; Zhang et al., 2023). However, while the use of MRI as a biomarker is well established, systematic evaluation of its test–retest reliability, particularly in the context of intervention-based longitudinal studies, remains scarce.

In this study, we assessed the validity of structural and functional MRI-derived brain features by quantifying both reproducibility and uncertainty. Specifically, we examined subcortical volume, cortical thickness, and functional connectivity within brain networks. Using test-retest data from 20 healthy participants, we calculated the repeatability coefficient to define the measurement uncertainty of each feature, and Pearson correlation coefficients to quantify reproducibility. These estimates then served as reference benchmarks to interpret changes in 24 patients undergoing 20 sessions of TMS. This dual-pronged approach allowed us to distinguish genuine treatment-related changes from natural biological and methodological variability.

Our test-retest analysis revealed that structural MRI measures, particularly subcortical volumes and cortical thickness, exhibited high reproducibility and low repeatability coefficients, indicating high reliability. Among these, the right caudate and left occipital cortex had the smallest RC%, while the left amygdala and left temporal pole had the largest variability among brain structures. These findings are consistent with previous reports demonstrating good reliability for cortical and subcortical volumetric measures, including gray and white matter, as well as thalamus volume (Heckendorf, Bakermans-Kranenburg, van Ijzendoorn, & Huffmeijer, 2019; Melzer et al., 2020). In contrast, functional connectivity features, including within-network connectivity across AAL and Yeo parcellations, demonstrated lower reproducibility and greater uncertainty. This pattern underscores the challenge of using resting-state fMRI in clinical research, where signal variability may be compounded by motion artifacts, scanner drift, or transient cognitive states (Botvinik-Nezer et al., 2020; Marek et al., 2022).

Interestingly, FA values from diffusion imaging demonstrated intermediate reliability, with variability influenced by region-specific susceptibility to noise, crossing fibers, and misalignment. While DTI-derived measures are often assumed to be reliable, our results emphasize the need for region-wise validation when interpreting FA changes. On the other hand, when evaluating changes following TMS intervention in patients, structural measures exhibited relatively minor alterations. Given that brain structure typically does not undergo significant modification over such a short period, these findings were expected.

Furthermore, understanding the direction of change in a parameter (i.e., whether it increases or decreases) can significantly aid the decision-making process. When the direction of change is of interest, it is important to note that an increase is indicated when the change in a measure exceeds the upper bound of CI for the RC, while a decrease is indicated when the change falls below the lower bound of the CI. With this approach, if an increase in a parameter is associated with a beneficial treatment response, patients exhibiting a decrease or no significant change in RC can be identified as non-responders, and treatment may be discontinued accordingly. Importantly, by incorporating both measurement uncertainty and directionality, clinicians can interpret neuroimaging changes at the individual level, rather than relying solely on group-level inferences. This strategy represents a valuable step toward advancing individualized, precision medicine.

The interpretability of individual patient changes also benefited from the use of RC confidence intervals derived from healthy controls. For instance, using the lower bound *RC*_*L*_ as a conservative threshold, we found that changes in FA in the cingulum bundle exceeded this uncertainty range in over 80% of patients, providing evidence for treatment-induced microstructural remodeling.

We found that anxiety was closely coupled with neural change: (i) greater reductions in anxiety were associated with larger within-network changes of the Yeo default mode network (DMN), and (ii) higher pre-treatment anxiety predicted greater cortical thickness change in the right frontal pole. Both effects survived Bonferroni correction and were observed in patients with severe anxiety. Our findings demonstrate that pre-treatment BAI scores may serve as a predictive marker for neural changes after applying TMS. The significant correlation between baseline anxiety levels and cortical thickness changes in the frontal pole suggests that individuals with higher pre-treatment anxiety exhibit greater structural plasticity in these regions. This particular region is also known as a critical node in emotion regulation and sensory processing, supporting the notion that anxiety levels may modulate neural responses to TMS (Roelofs, Bramson, & Toni, 2023; Sawalha et al., 2021; H. Y. Wang et al., 2018). Clinically, baseline anxiety might index a phenotype with greater capacity for treatment-related plasticity, suggesting that anxiety measures could help identify patients who are more likely to exhibit measurable brain change and, potentially, to benefit from network-informed modulation strategies.

Our findings indicated that TMS treatment significantly reduced both depression and anxiety symptoms, regardless of the participant’s gender. While the reduction in depressive symptoms did not differ significantly between men and women, the improvement in anxiety symptoms was influenced by gender, with women showing a greater reduction in anxiety levels. This does not suggest that the treatment was ineffective for one group, but rather that its efficacy varied across groups. This result is consistent with previous studies, which have reported that females tend to be more responsive to TMS interventions (Adamson et al., 2022; Hanlon & McCalley, 2022). Several factors may contribute to this difference, including a shorter distance between the skull and the cerebral cortex, higher gray matter density in prefrontal regions, and elevated estradiol levels in women (Hanlon & McCalley, 2022).

We also observed a strong positive association between change in anxiety and change in within-network connectivity of the Yeo DMN (r=0.78, p=0.001): participants with smaller reductions in anxiety showed greater increases in DMN within-network connectivity, whereas greater anxiety relief was accompanied by decreases (or minimal increases) in DMN cohesion. This pattern suggests that symptom improvement may track a normalization of DMN dynamics implicated in self-referential processing, particularly in patients with severe anxiety.

Future studies should extend this methodology to larger and more diverse samples, incorporate additional MRI modalities such as task-based fMRI, and evaluate clinical outcomes longitudinally. Collectively, this work establishes a foundation for quantifying and interpreting MRI-based biomarkers in real-world clinical contexts, with direct implications for individualized treatment planning in psychiatry.

## Data Availability

All data produced in the present study are available upon reasonable request to the authors

## Notes

### Competing Interest Statement

The authors have declared no competing interest.

### Author Declarations

All participants provided informed consent according to the study protocol approved by the ethics committee of research, Iran University of Medical Sciences (Ethic code: IR.IUMS.REC.1399.1419).

