## Supplementary for "Assessing MRI Biomarker Repeatability to Guide Individualized TMS Treatment in Psychiatry"

**Supplementary Material**

This document provides additional information and results supporting the findings presented in the main manuscript. The following materials are included:

Figure S1. Test-retest reliability of MRI measures

Figure S2. Association between baseline depression severity (BDI-pre) and changes in MRI measures, stratified by mild and severe depression groups.

Figure S3. Association between baseline anxiety severity (BAI-pre) and changes in MRI measures, stratified by mild and severe anxiety groups.

Figure S4. Association between changes in depression scores (ΔBDI) and changes in MRI measures, stratified by mild and severe depression groups.

Figure S5. Association between changes in anxiety scores (ΔBAI) and changes in MRI measures, stratified by mild and severe anxiety groups.

Table S1. Brain regions and network assignments: Detailed information including region names, stereotaxic coordinates (x, y, z), and network labels based on AAL and Yeo atlases.

Table S2. Repeatability coefficient (RC) estimates for MRI measures.

The following figures display box plots comparing MRI measures between test and retest sessions for healthy controls. For each measure, green boxes represent test session data and blue boxes represent retest session data. The horizontal line within each box indicates the median. Paired t-tests were performed to assess systematic differences between sessions.

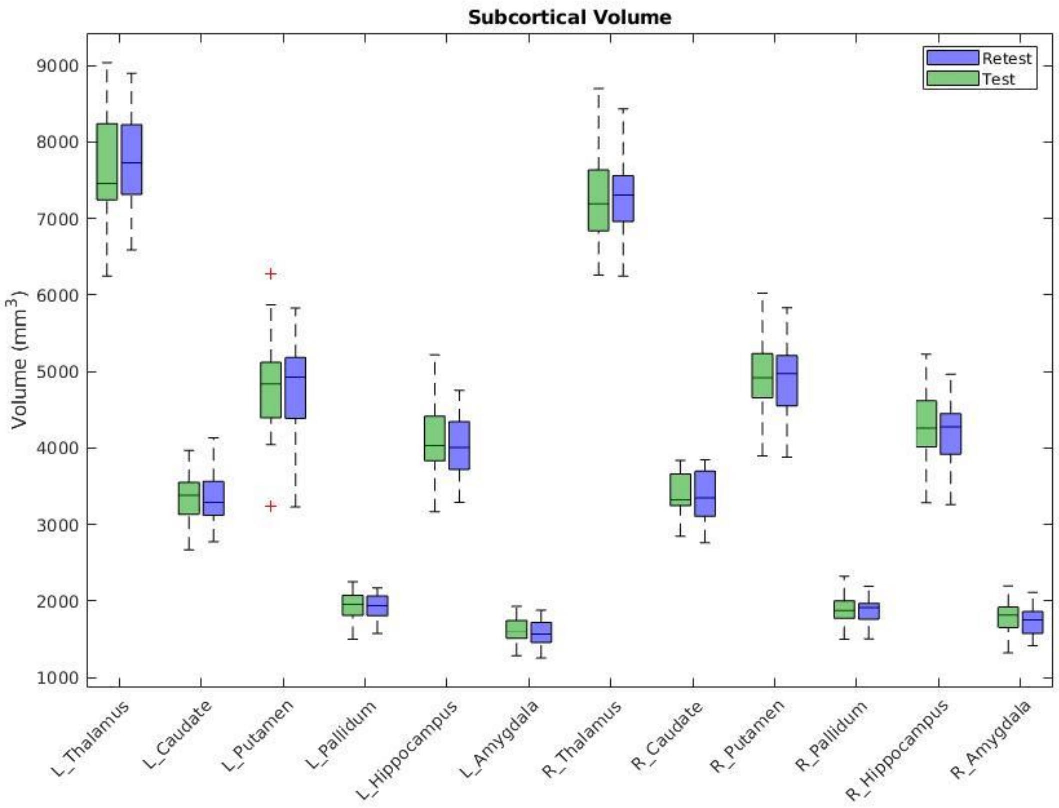


(a)

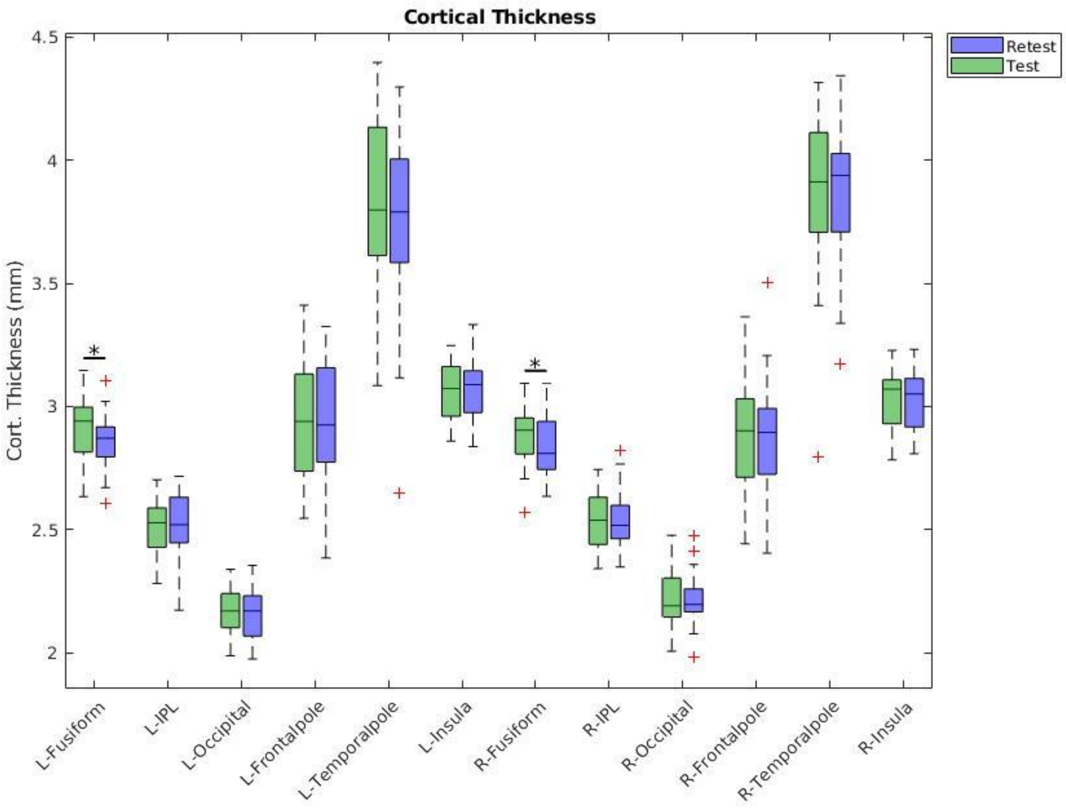
Cortical Thickness
Retest
Test
Cort. Thickness (mm)
4.5
4
3.5
3
2.5
2
*
*
L-Fusiform
L-IPL
L-Occipital
L-Frontalpole
L-Temporalpole
L-Insula
R-Fusiform
R-IPL
R-Occipital
R-Frontalpole
R-Temporalpole
R-Insula

(b)

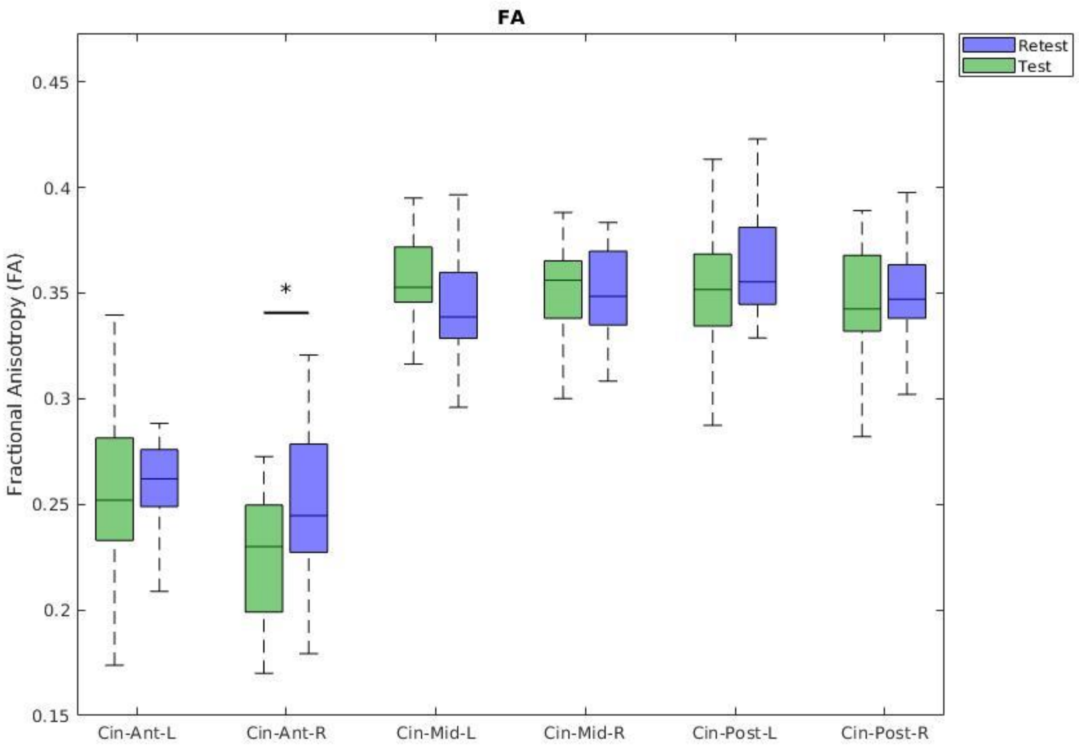
FA
Retest
Test
Fractional Anisotropy (FA)
0.45
0.4
0.35
0.3
0.25
0.2
0.15
*
Cin-Ant-L
Cin-Ant-R
Cin-Mid-L
Cin-Mid-R
Cin-Post-L
Cin-Post-R

(c)

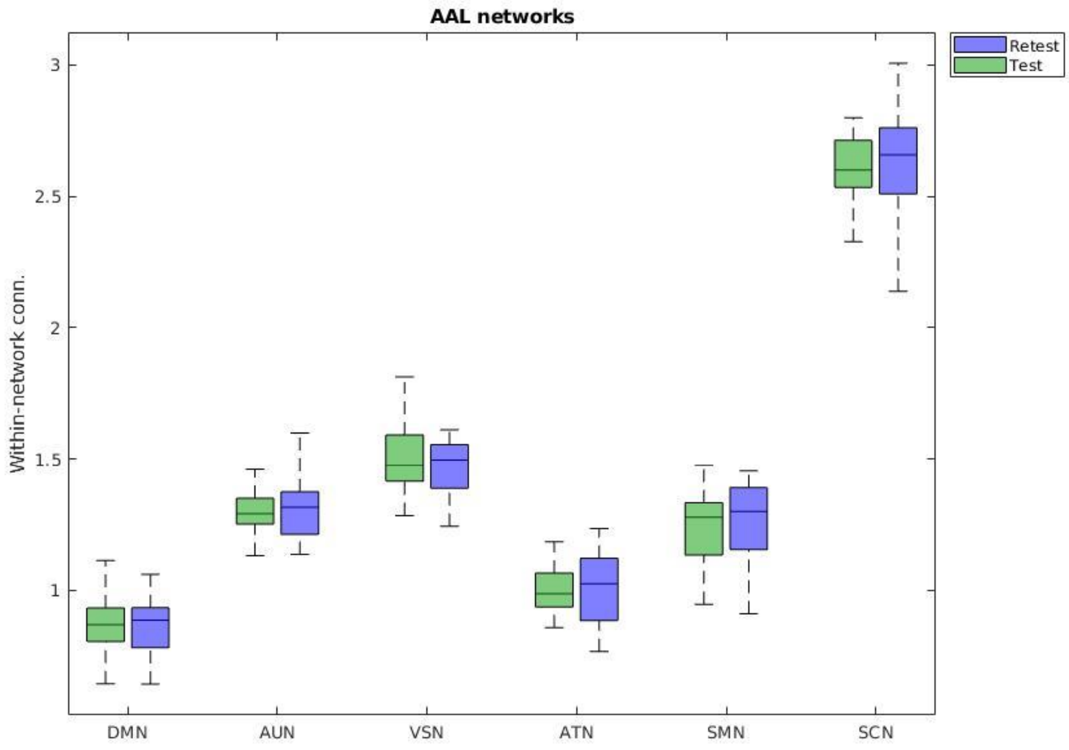
AAL networks
Retest
Test
Within-network conn.
3
2.5
2
1.5
1
DMN
AUN
VSN
ATN
SMN
SCN

(d)

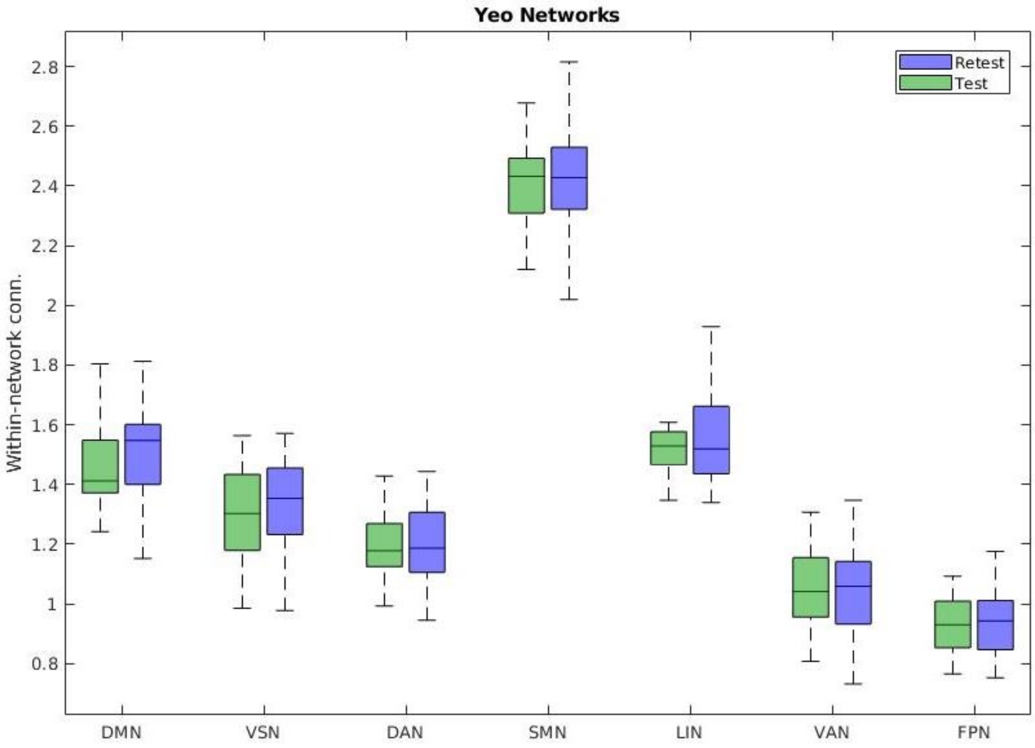


(e)

**Figure S1**. Box plot of a) subcortical volume, b) cortical thickness, c) FA, d) within-network connectivity of AAL networks, and e) within-network connectivity of Yeo networks for test (green boxes) and retest (blue boxes) data. The median is indicated by a line across the box. A paired t-test showed no differences between test and retest data except for the thickness of the right Frontal pole and right anterior Cingulum. (* $p<0.05$) L, left; R, right; IPL, inferior parietal lobule; DMN, default mode network; VSN, visual network; DAN, dorsal attention network; FPN, fronto-parietal network; SMN, sensorimotor network; LIN, limbic network; AUN, auditory network; ATN, attention network; SCN, subcortical network.

The following figures display correlation analyses between pre-treatment depression scores (BDI-pre) and changes in structural and functional MRI indices. Patients were stratified into two groups based on baseline symptom severity: mild depression (BDI-pre < 20) and severe depression (BDI-pre ≥ 20). For each group, correlations are shown for five MRI measures: (a) subcortical volumes, (b) cortical thickness, (c) fractional anisotropy (FA), (d) within-network functional connectivity based on the AAL atlas, and (e) within-network functional connectivity based on the Yeo atlas.

### 1- Mild depression group (BDI < 20)

#### a) Subcortical volume

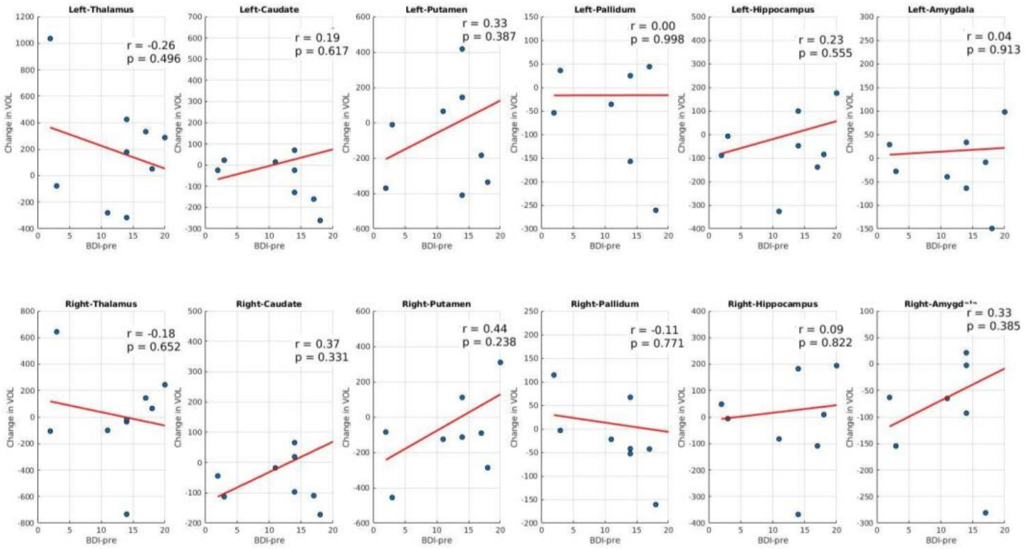


#### b) Cortical Thickness

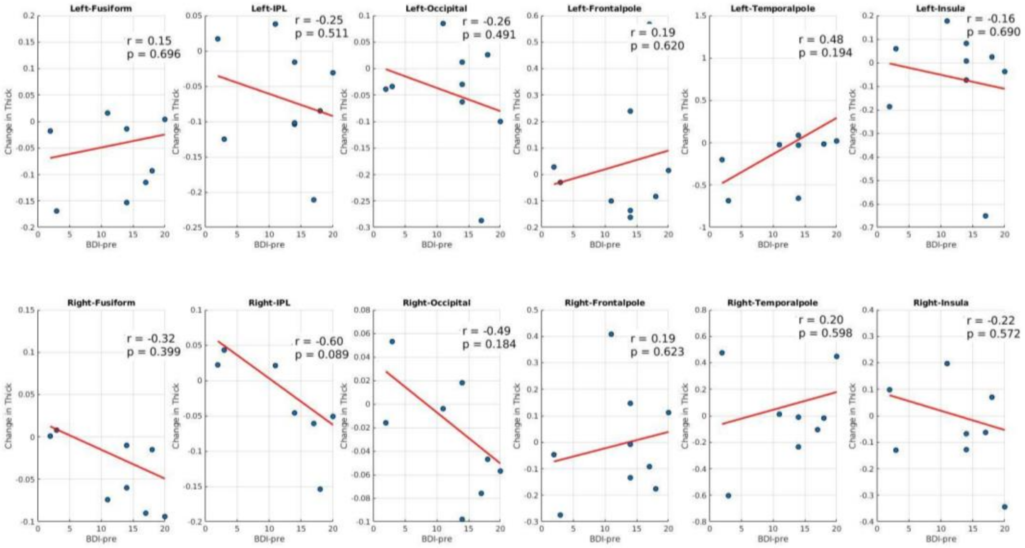

c) **FA**

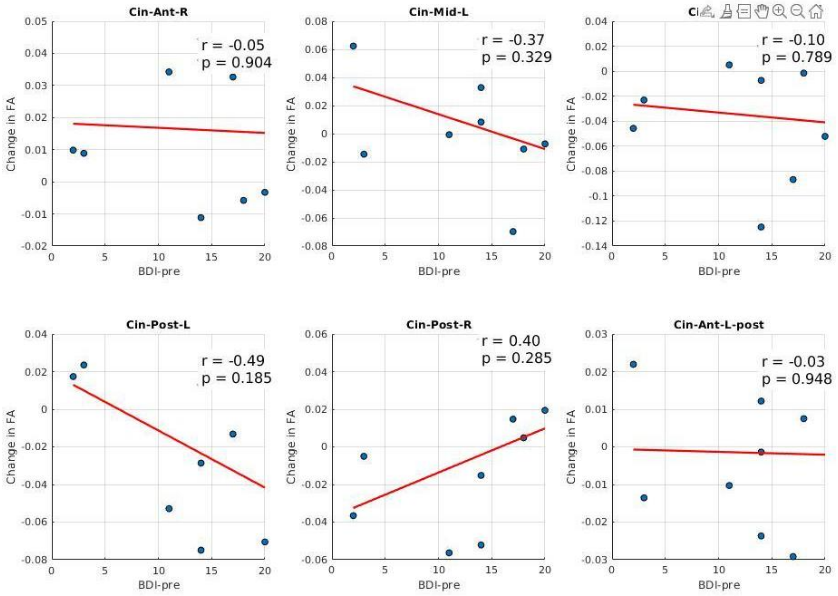


**d) AAL**

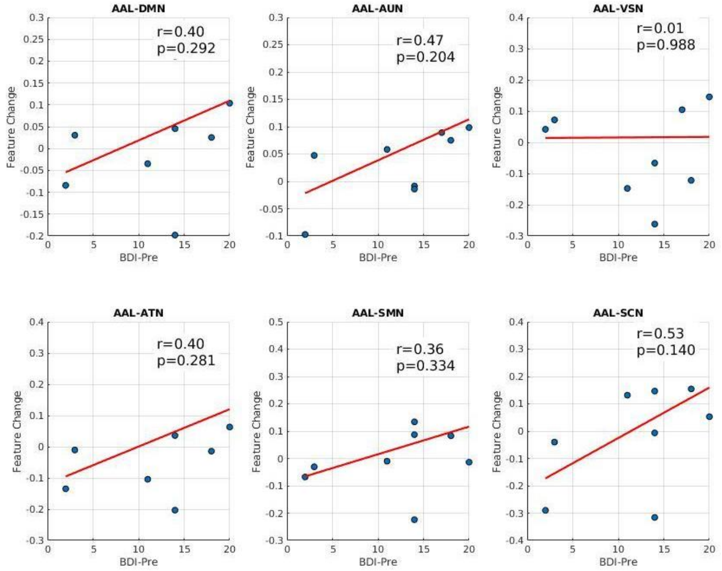


e) **Yeo**

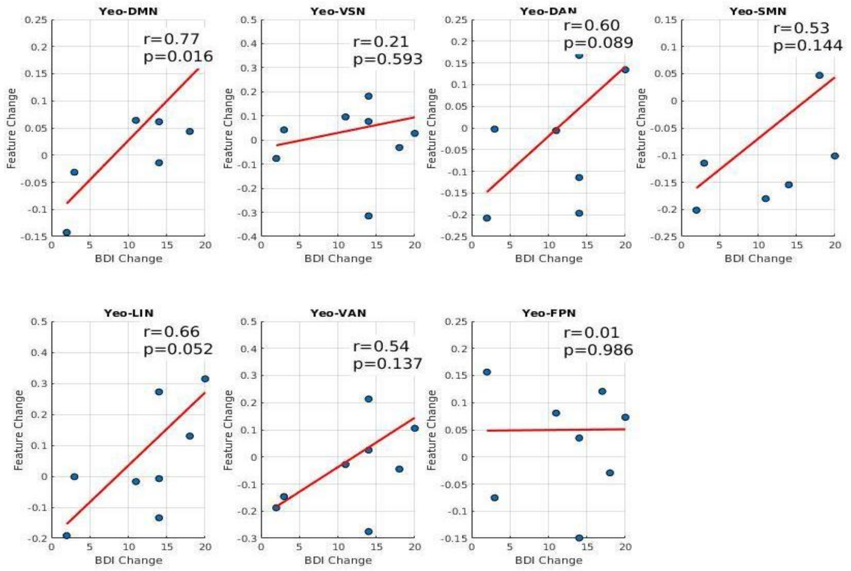


### 2- Severe depression (BDI > 20)

#### a) Subcortical Volume

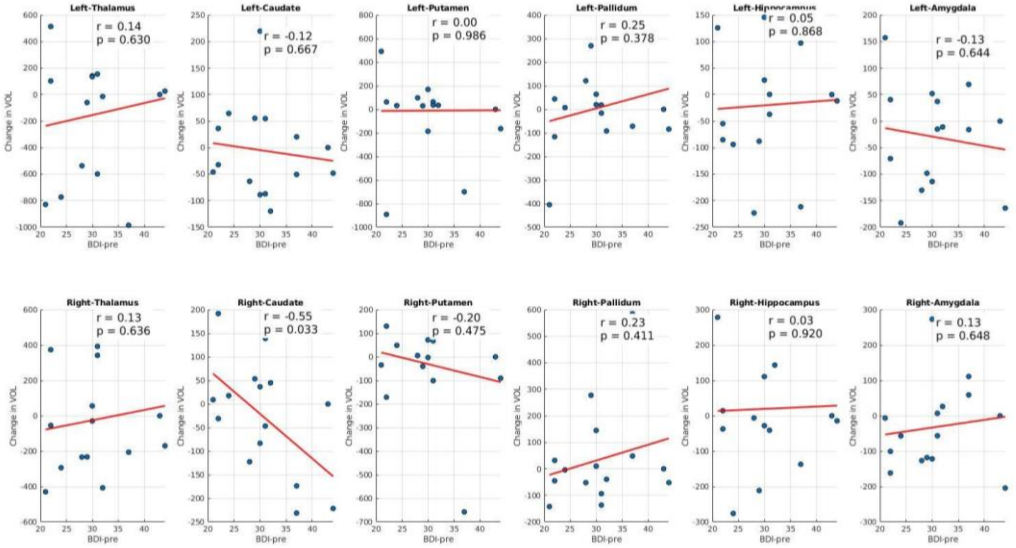


#### b) Cortical Thickness

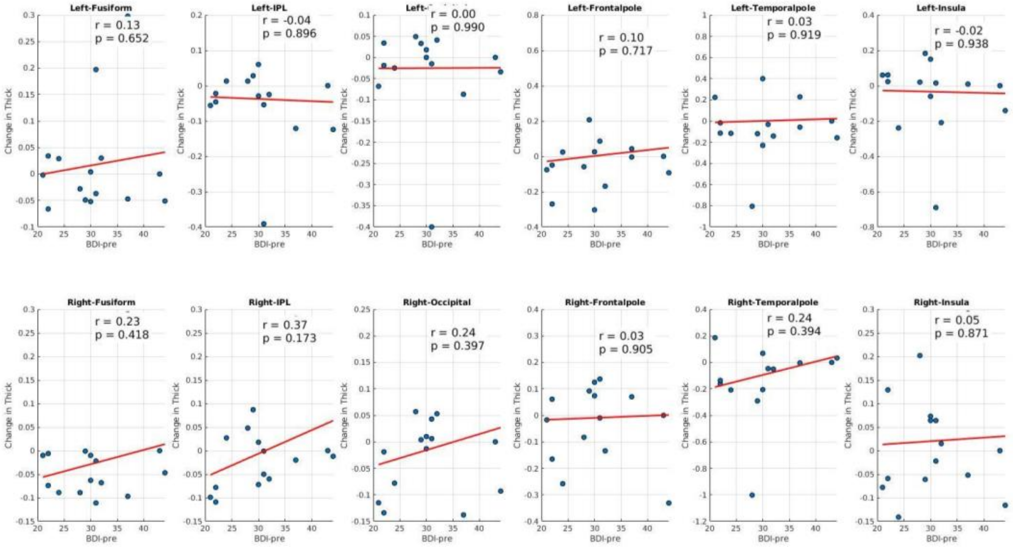
Left-Fusiform
r = 0.13
p = 0.652
Left-IPL
r = -0.04
p = 0.896
r = 0.00
p = 0.990
Left-Frontalpole
r = 0.10
p = 0.717
Left-Temporalpole
r = 0.03
p = 0.919
Left-Insula
r = -0.02
p = 0.938
Right-Fusiform
r = 0.23
p = 0.418
Right-IPL
r = 0.37
p = 0.173
Right-Occipital
r = 0.24
p = 0.397
Right-Frontalpole
r = 0.03
p = 0.905
Right-Temporalpole
r = 0.24
p = 0.394
Right-Insula
r = 0.05
p = 0.871
Change in Thick
BDI-pre

## c) FA

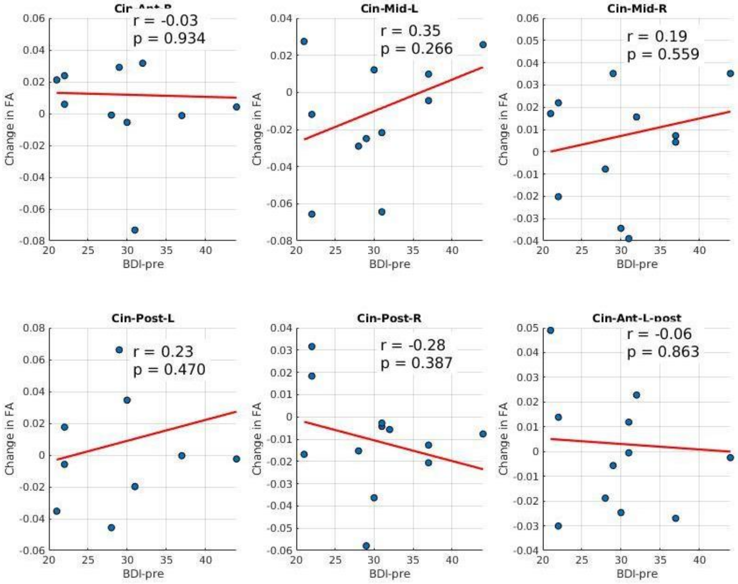
r = -0.03
p = 0.934
Cin-Mid-L
r = 0.35
p = 0.266
Cin-Mid-R
r = 0.19
p = 0.559
Cin-Post-L
r = 0.23
p = 0.470
Cin-Post-R
r = -0.28
p = 0.387
Cin-Ant-L-post
r = -0.06
p = 0.863
Change in FA
BDI-pre

### d) AAL

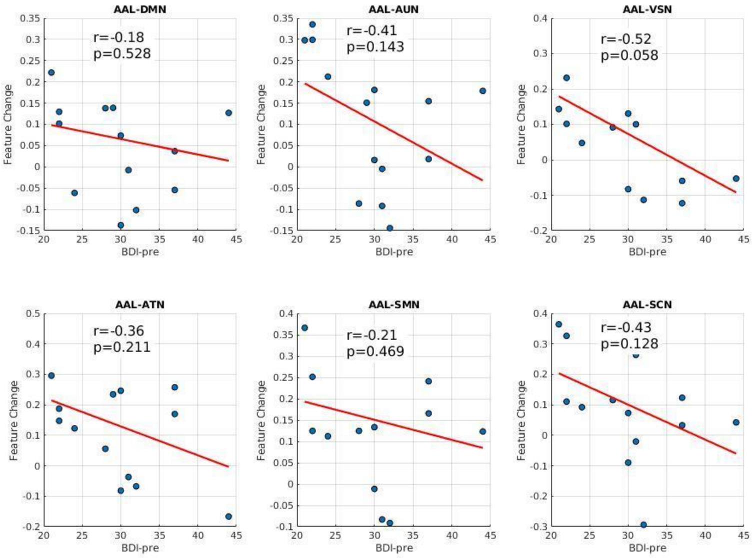


**e) Yeo**

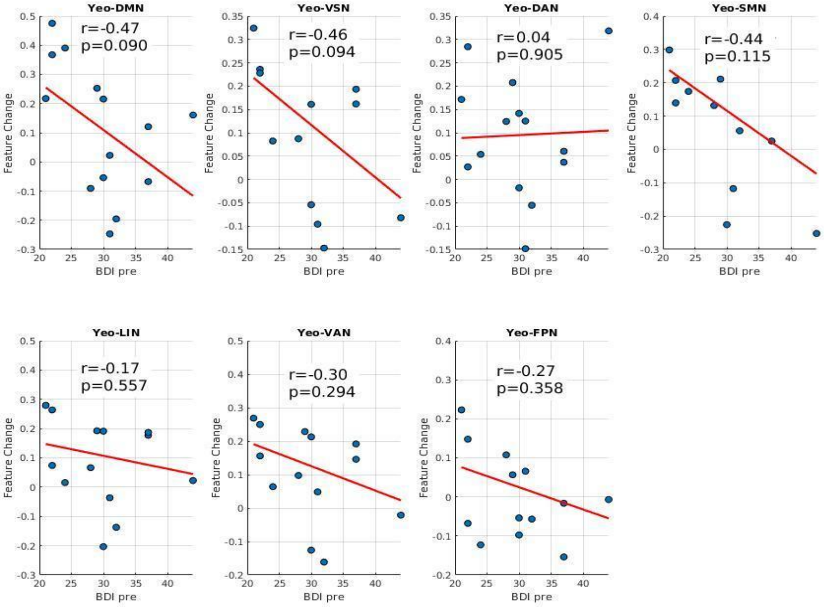


Figure S2. Association between baseline depression severity and changes in morphometric and connectivity MRI measures. This figure displays correlation coefficients (r) between pre-treatment depression scores (BDI-pre) and changes (Δ) in structural and functional MRI indices across two patient subgroups stratified by baseline symptom severity. Positive and negative values indicate direct and inverse correlations, respectively.

(1) Mild depression group (BDI-pre < 20): Correlation coefficients between BDI-pre and changes in MRI measures for patients with mild baseline symptoms: (a) Subcortical volumes; (b) Cortical thickness; (c) Fractional anisotropy (FA); (d) Within-network functional connectivity based on AAL atlas; (e) Within-network functional connectivity based on Yeo atlas.

(2) Severe depression group (BDI-pre ≥ 20): Correlation coefficients between BDI-pre and changes in MRI measures for patients with severe baseline symptoms: (a) Subcortical volumes; (b) Cortical thickness; (c) Fractional anisotropy (FA); (d) Within-network functional connectivity based on AAL atlas; (e) Within-network functional connectivity based on Yeo atlas.

The following figures display correlation analyses between pre-treatment anxiety scores (BAI-pre) and changes in structural and functional MRI indices. Patients were stratified into two groups based on baseline symptom severity: mild anxiety (BAI-pre < 15) and severe anxiety (BAI-pre ≥ 15). For each group, correlations are shown for five MRI measures: (a) subcortical volumes, (b) cortical thickness, (c) fractional anisotropy (FA), (d) within-network functional connectivity based on the AAL atlas, and (e) within-network functional connectivity based on the Yeo atlas.

### 1- Mild anxiety group (BAI < 15)

#### a) Subcortical Volume

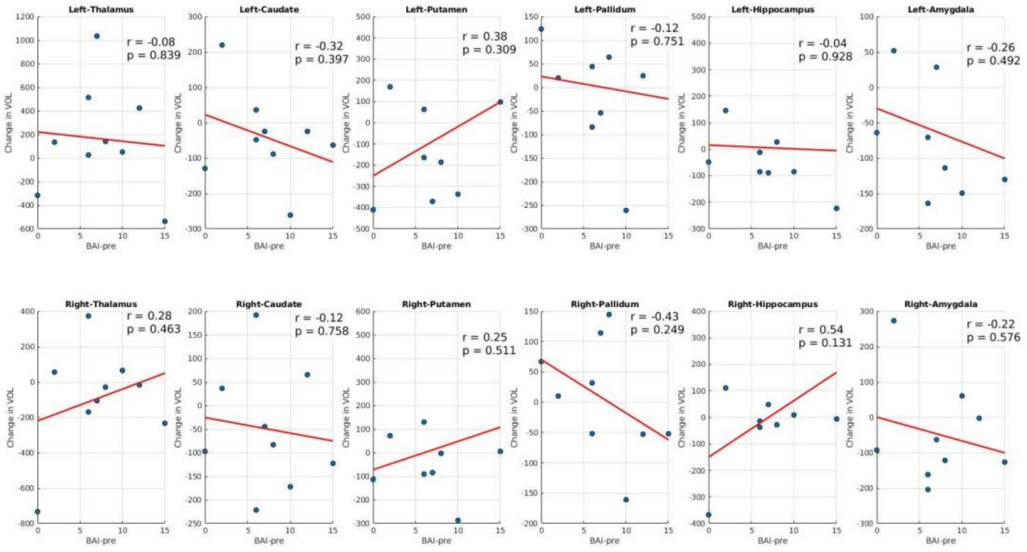


#### b) Cortical Thickness

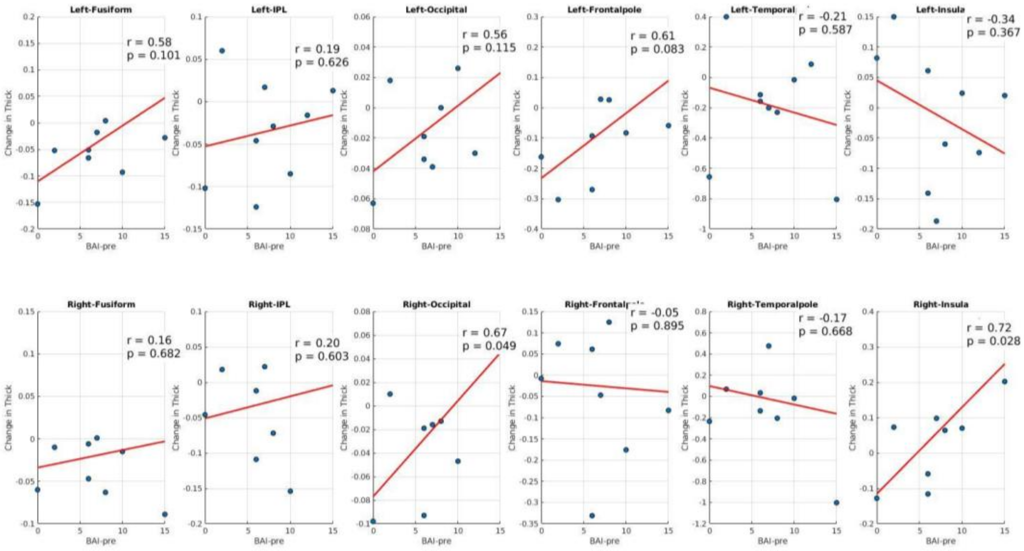

Left-Fusiform
r = 0.58
p = 0.101
Left-IPL
r = 0.19
p = 0.626
Left-Occipital
r = 0.56
p = 0.115
Left-Frontalpole
r = 0.61
p = 0.083
Left-Temporal
r = -0.21
p = 0.587
Left-Insula
r = -0.34
p = 0.367
Right-Fusiform
r = 0.16
p = 0.682
Right-IPL
r = 0.20
p = 0.603
Right-Occipital
r = 0.67
p = 0.049
r = -0.05
p = 0.895
Right-Temporalpole
r = -0.17
p = 0.668
Right-Insula
r = 0.72
p = 0.028
Change in Thick
BAI-pre


## c) FA

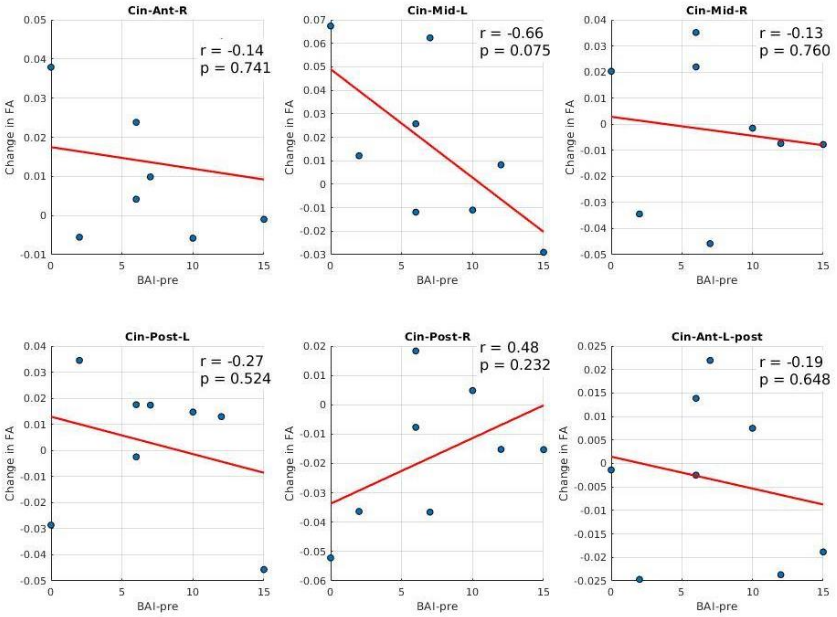

Cin-Ant-R
r = -0.14
p = 0.741
Cin-Mid-L
r = -0.66
p = 0.075
Cin-Mid-R
r = -0.13
p = 0.760
Cin-Post-L
r = -0.27
p = 0.524
Cin-Post-R
r = 0.48
p = 0.232
Cin-Ant-L-post
r = -0.19
p = 0.648
Change in FA
BAI-pre


### d) AAL

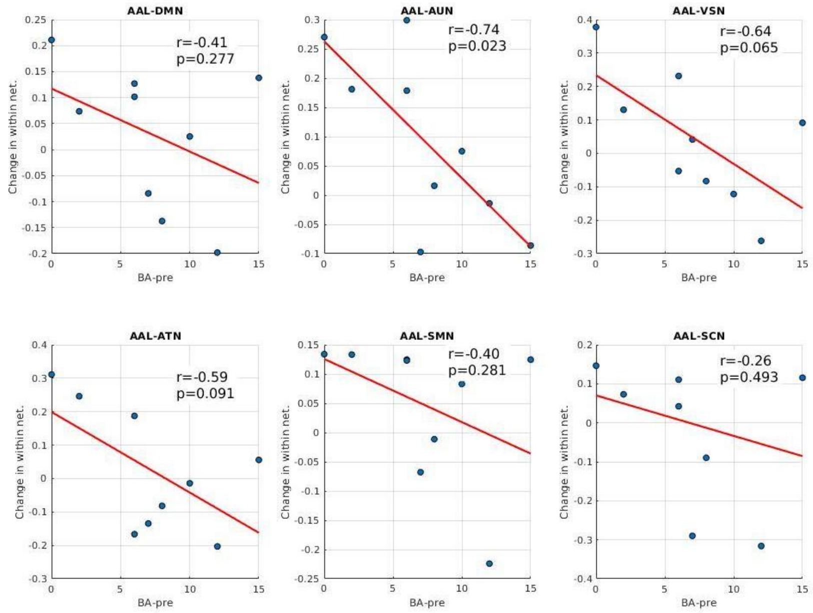


**e) Yeo**

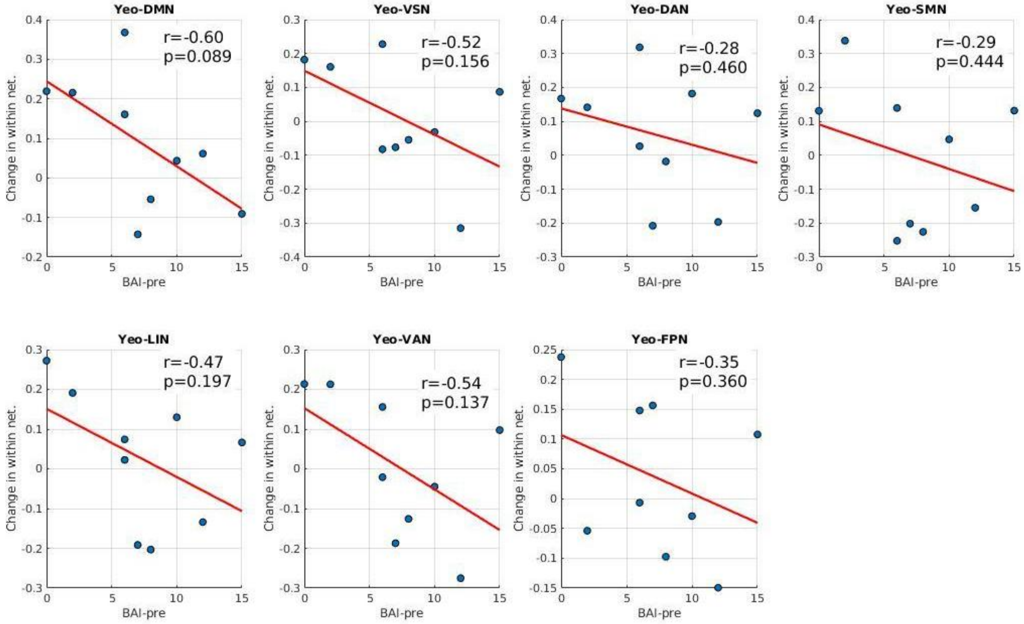


2- Severe anxiety group (BAI > 15)

**a) Subcortical Volume**

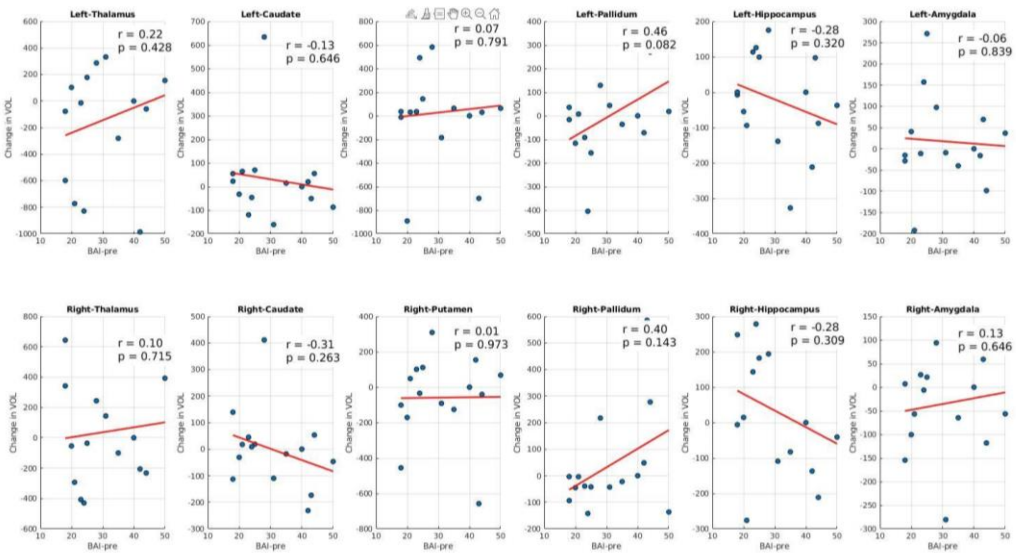


### b) Cortical Thickness

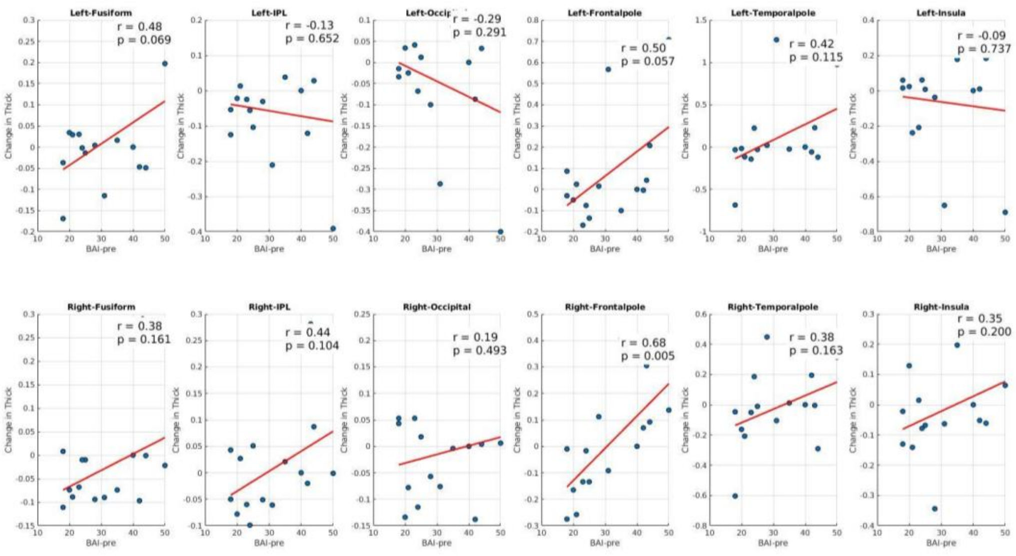


## c) FA

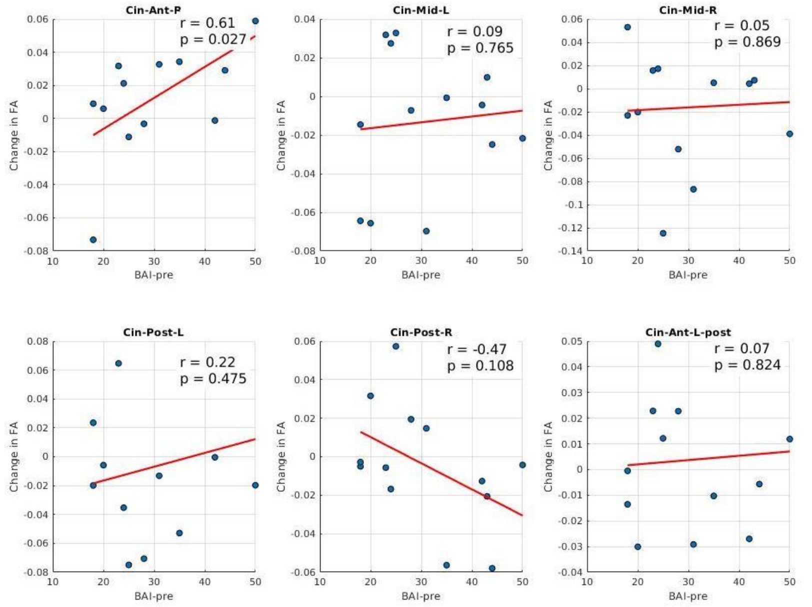

Cin-Ant-P
r = 0.61
p = 0.027
Cin-Mid-L
r = 0.09
p = 0.765
Cin-Mid-R
r = 0.05
p = 0.869
Cin-Post-L
r = 0.22
p = 0.475
Cin-Post-R
r = -0.47
p = 0.108
Cin-Ant-L-post
r = 0.07
p = 0.824
Change in FA
BAI-pre


### d) AAL

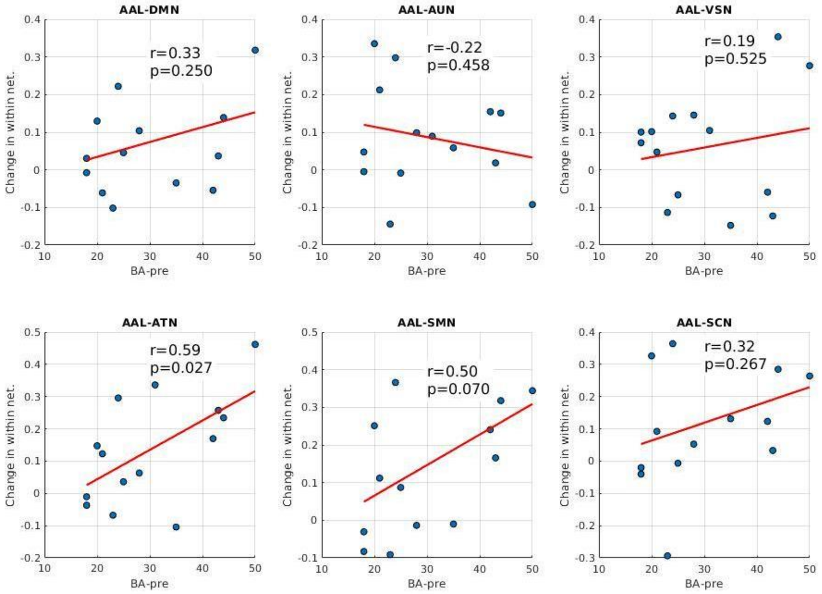

AAL-DMN
r=0.33
p=0.250
AAL-AUN
r=-0.22
p=0.458
AAL-VSN
r=0.19
p=0.525
AAL-ATN
r=0.59
p=0.027
AAL-SMN
r=0.50
p=0.070
AAL-SCN
r=0.32
p=0.267
Change in within net.
BA-pre


### e) Yeo

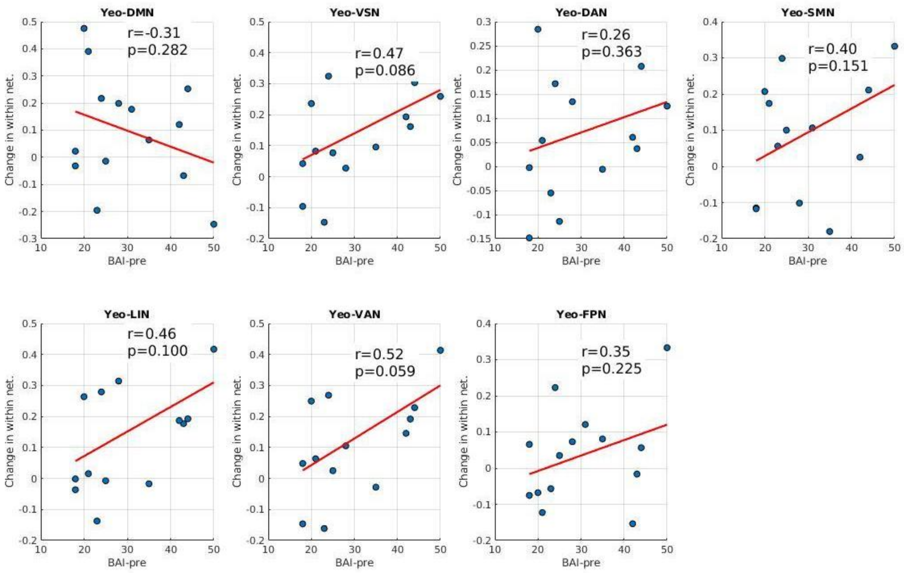


Figure S3. Association between baseline anxiety severity and changes in morphometric and connectivity MRI measures. This figure displays correlation coefficients (r) between pre-treatment anxiety scores (BAI-pre) and changes (Δ) in structural and functional MRI indices across two patient subgroups stratified by baseline symptom severity. Positive and negative values indicate direct and inverse correlations, respectively. (1) Mild anxiety group (BAI-pre < 15): Correlation coefficients between BAI-pre and changes in MRI measures for patients with mild baseline anxiety symptoms: (a) Subcortical volumes; (b) Cortical thickness; (c) Fractional anisotropy (FA); (d) Within-network functional connectivity based on AAL atlas; (e) Within-network functional connectivity based on Yeo atlas. (2) Severe anxiety group (BAI-pre ≥ 15): Correlation coefficients between BAI-pre and changes in MRI measures for patients with severe baseline anxiety symptoms: (a) Subcortical volumes; (b) Cortical thickness; (c) Fractional anisotropy (FA); (d) Within-network functional connectivity based on AAL atlas; (e) Within-network functional connectivity based on Yeo atlas.

The following figures display correlation analyses between changes in depression scores and changes in structural and functional MRI indices. Patients were stratified into two groups based on baseline symptom severity: mild depression (BDI-pre < 20) and severe depression (BDI-pre ≥ 20). For each group, correlations are shown for five MRI measures: (a) subcortical volumes, (b) cortical thickness, (c) fractional anisotropy (FA), (d) within-network functional connectivity based on the AAL atlas, and (e) within-network functional connectivity based on the Yeo atlas.

1- Mild depression group (BDI <= 20)

**a) Subcortical volume**

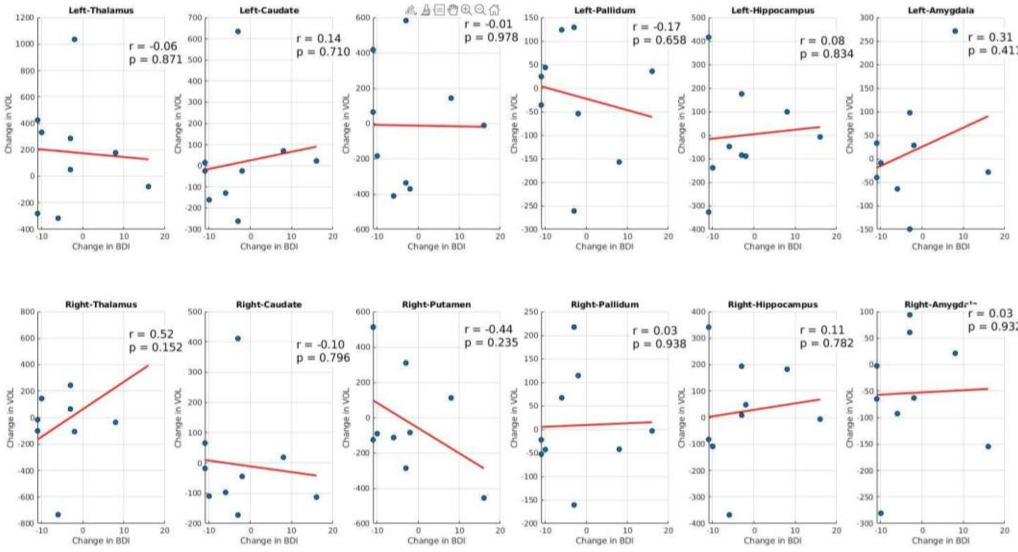


### b) Cortical Thickness

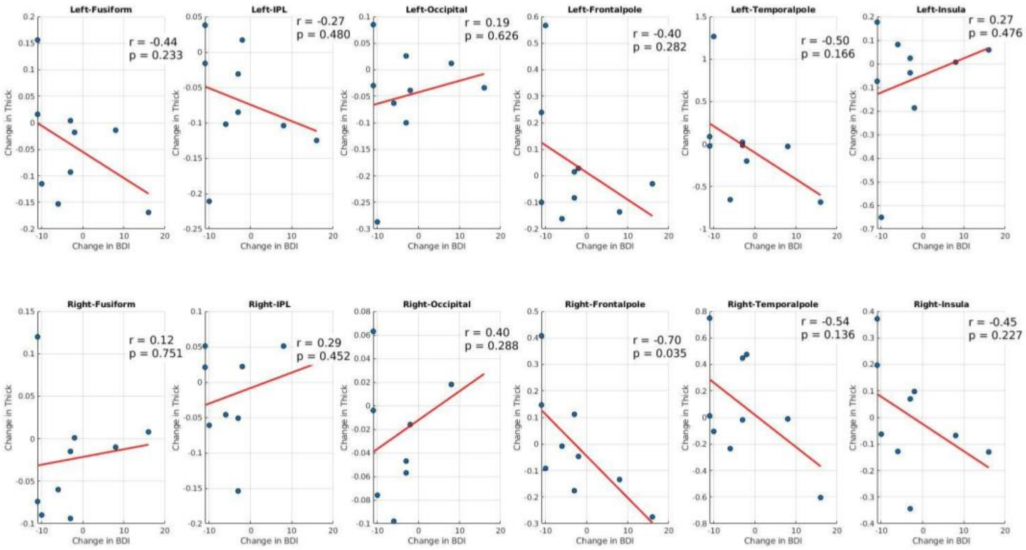


## c) FA

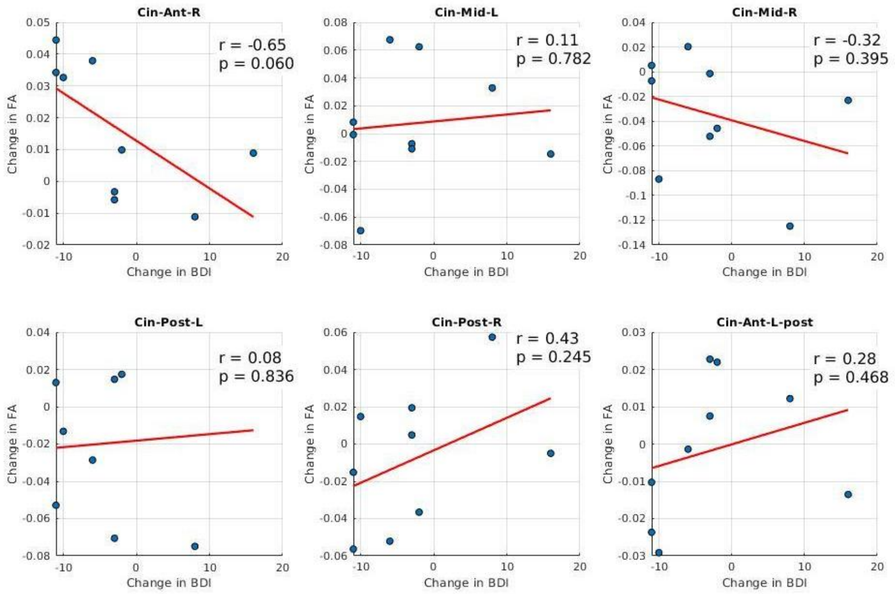
Cin-Ant-R
r = -0.65
p = 0.060
Cin-Mid-L
r = 0.11
p = 0.782
Cin-Mid-R
r = -0.32
p = 0.395
Cin-Post-L
r = 0.08
p = 0.836
Cin-Post-R
r = 0.43
p = 0.245
Cin-Ant-L-post
r = 0.28
p = 0.468
Change in FA
Change in BDI

### d) AAL

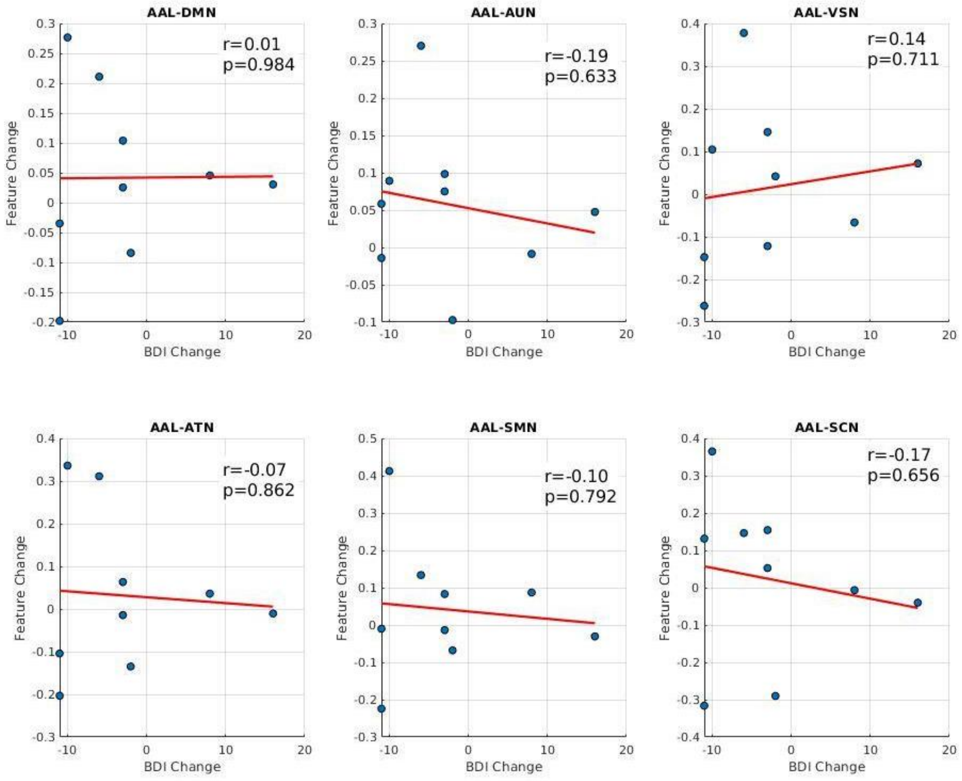
AAL-DMN
r=0.01
p=0.984
AAL-AUN
r=-0.19
p=0.633
AAL-VSN
r=0.14
p=0.711
AAL-ATN
r=-0.07
p=0.862
AAL-SMN
r=-0.10
p=0.792
AAL-SCN
r=-0.17
p=0.656
Feature Change
BDI Change

### e) **Yeo**

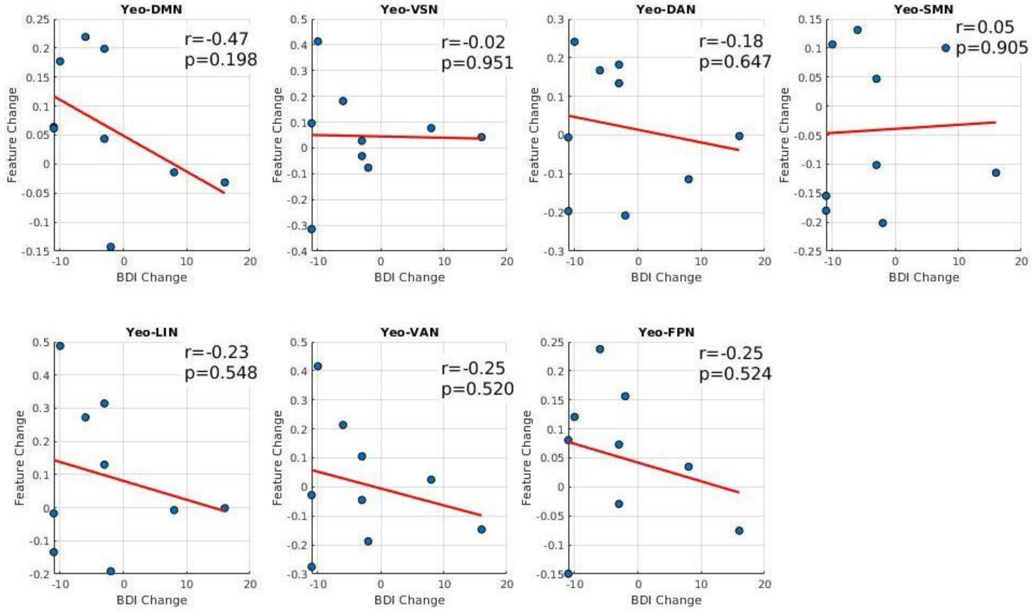


### 2- Severe depression group (BDI > 20)

#### a) Subcortical volume

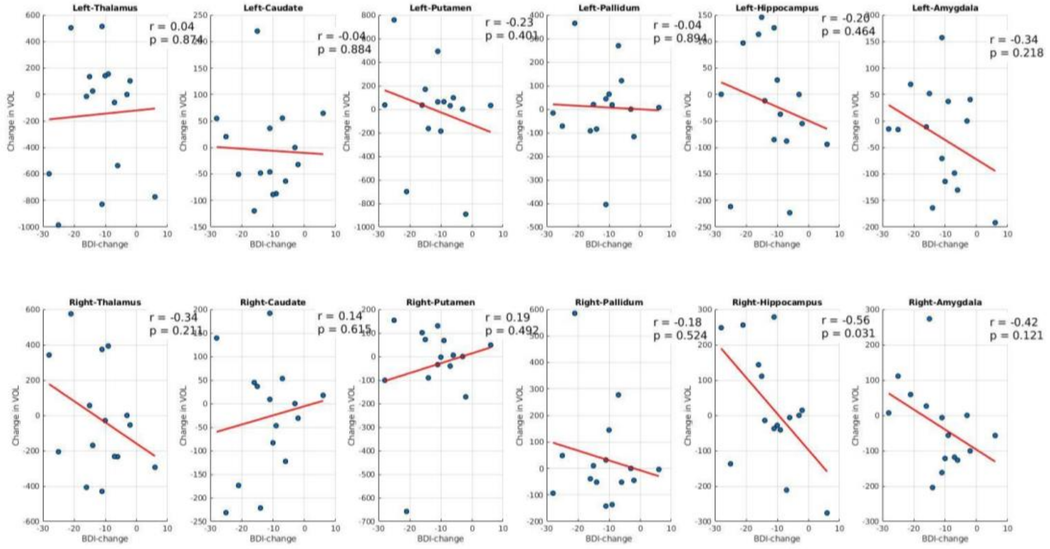


#### b) Cortical Thickness

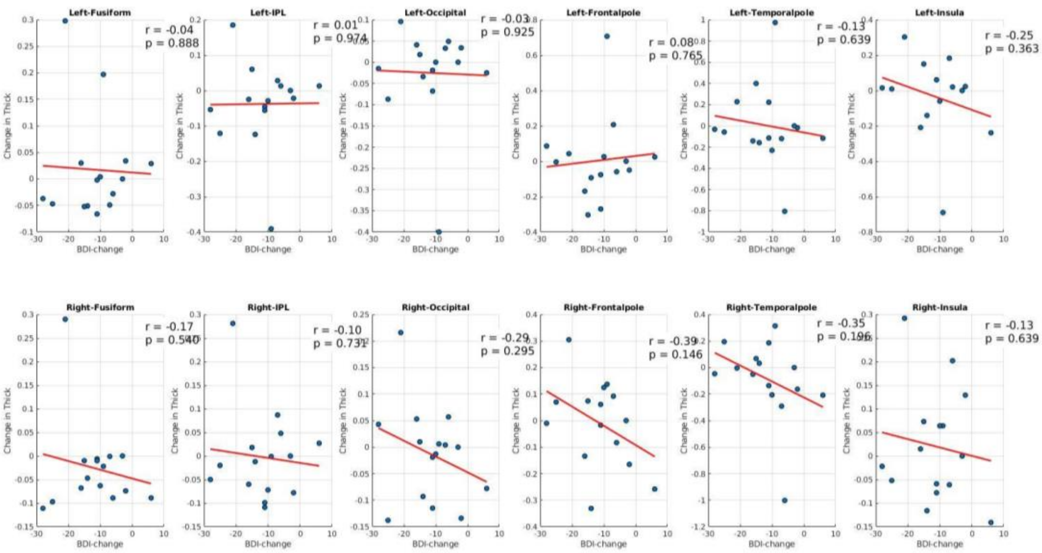
Left-Fusiform
r = -0.04
p = 0.888
Left-IPL
Left-Occipital
r = -0.03
p = 0.925
Left-Frontalpole
Left-Temporalpole
r = -0.13
p = 0.639
Left-Insula
r = -0.25
p = 0.363
Right-Fusiform
Right-IPL
Right-Occipital
Right-Frontalpole
Right-Temporalpole
Right-Insula
r = -0.13
p = 0.639
Change in Thick
BDI-change

**c) FA**

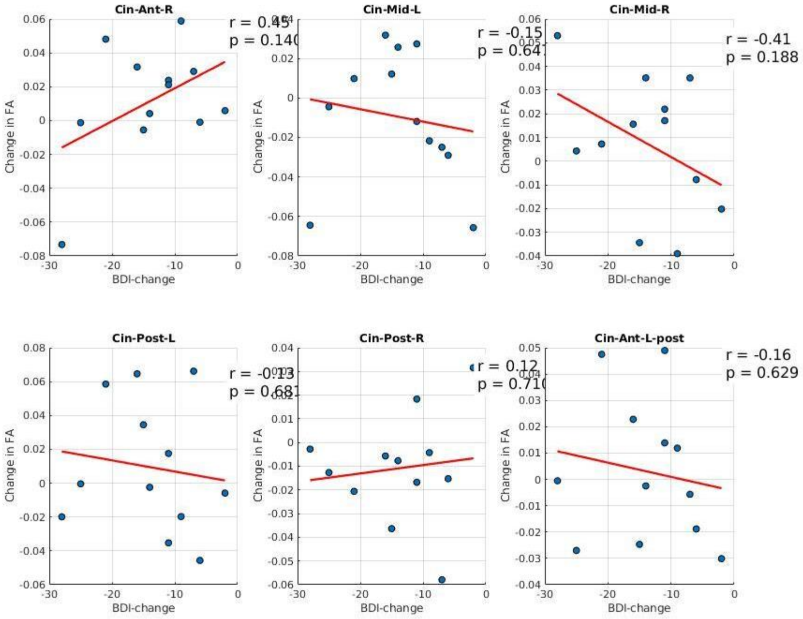
Cin-Ant-R
Cin-Mid-L
Cin-Mid-R
r = -0.41
p = 0.188
Cin-Post-L
Cin-Post-R
Cin-Ant-L-post
r = -0.16
p = 0.629
Change in FA
BDI-change

**d) AAL**

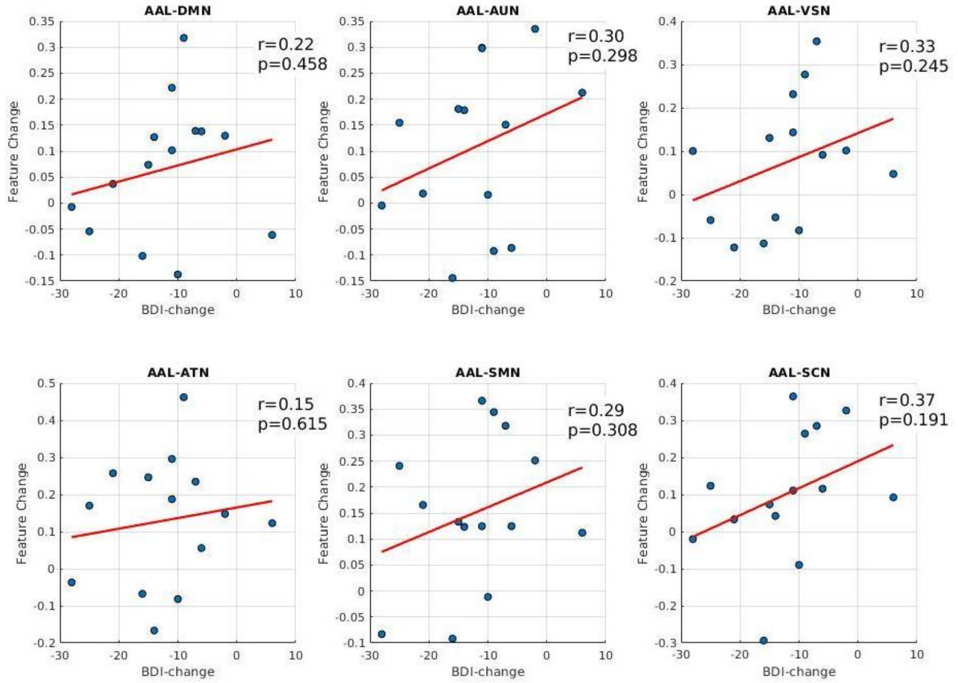


**e) Yeo**

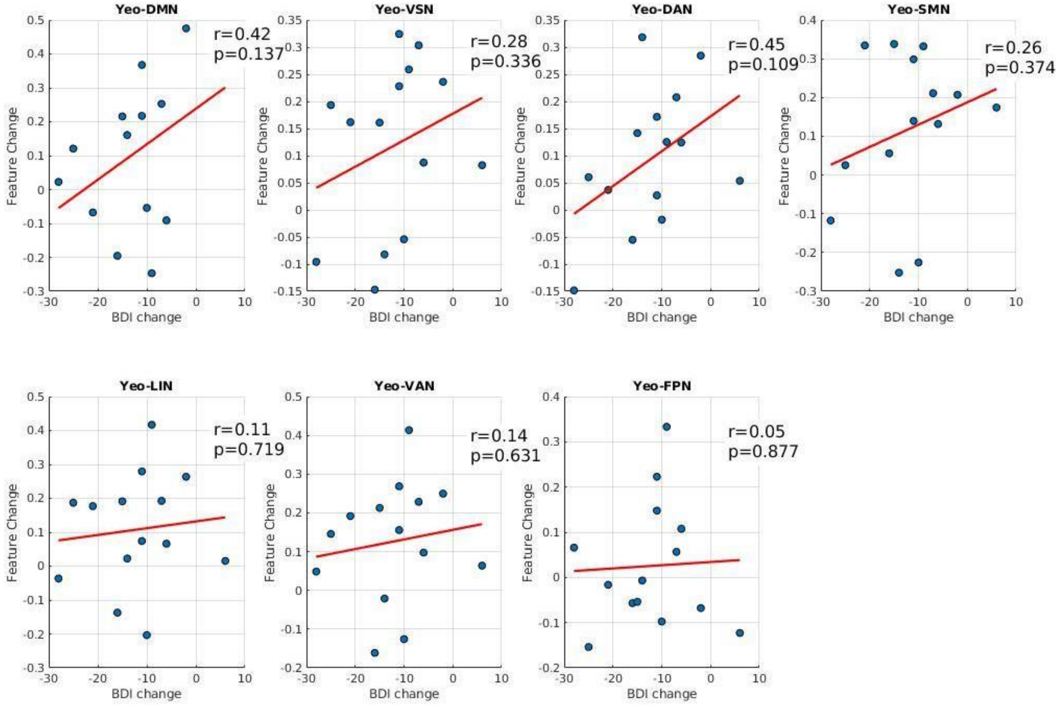


**Figure S4.** Association between changes in depression scores and changes in MRI measures. This figure displays correlation coefficients (r) between changes in depression scores (BDI-post - BDI-pre) and changes in structural and functional MRI indices across two patient subgroups stratified by baseline symptom severity. Negative ΔBDI values indicate improvement in depressive symptoms. Positive and negative correlation coefficients indicate direct and inverse associations, respectively. (1) Mild depression group (BDI-pre < 20): Correlation coefficients between BDI changes and changes in MRI measures for patients with mild baseline symptoms: (a) Subcortical volumes; (b) Cortical thickness; (c) Fractional anisotropy (FA); (d) Within-network functional connectivity based on AAL atlas; (e) Within-network functional connectivity based on Yeo atlas. (2) Severe depression group (BDI-pre ≥ 20): Correlation coefficients between ΔBDI and changes in MRI measures for patients with severe baseline symptoms: (a) Subcortical volumes; (b) Cortical thickness; (c) Fractional anisotropy (FA); (d) Within-network functional connectivity based on AAL atlas; (e) Within-network functional connectivity based on Yeo atlas.

The following figures display correlation analyses between changes in anxiety scores (ΔBAI = BAI-post - BAI-pre) and changes in structural and functional MRI indices. Patients were stratified into two groups based on baseline symptom severity: mild anxiety (BAI-pre < 15) and severe anxiety (BAI-pre ≥ 15). For each group, correlations are shown for five MRI measures: (a) subcortical volumes, (b) cortical thickness, (c) fractional anisotropy (FA), (d) within-network functional connectivity based on the AAL atlas, and (e) within-network functional connectivity based on the Yeo atlas.

### 1- Mild anxiety (BAI < 15): 9

#### a) Subcortical volume

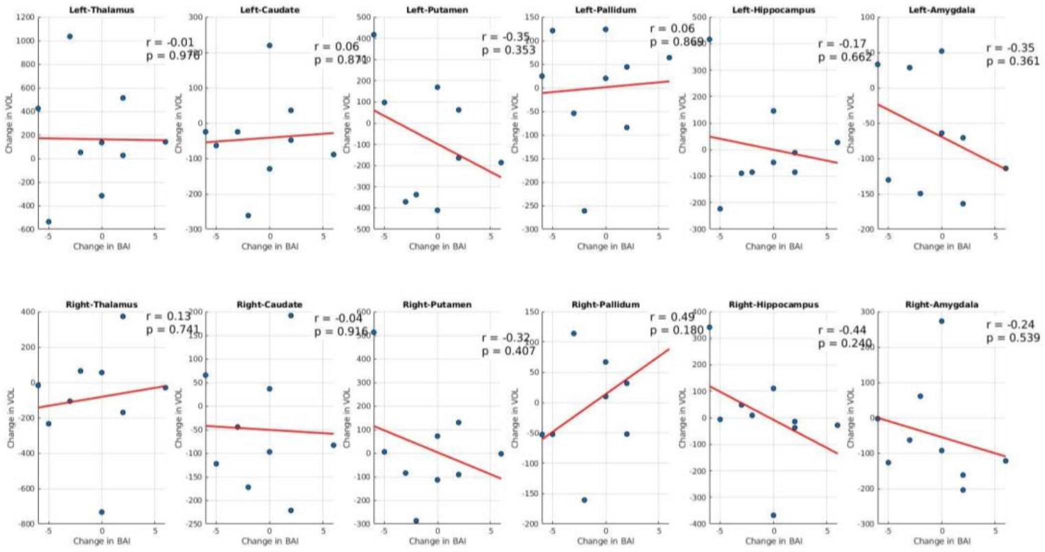


#### b) Cortical Thickness

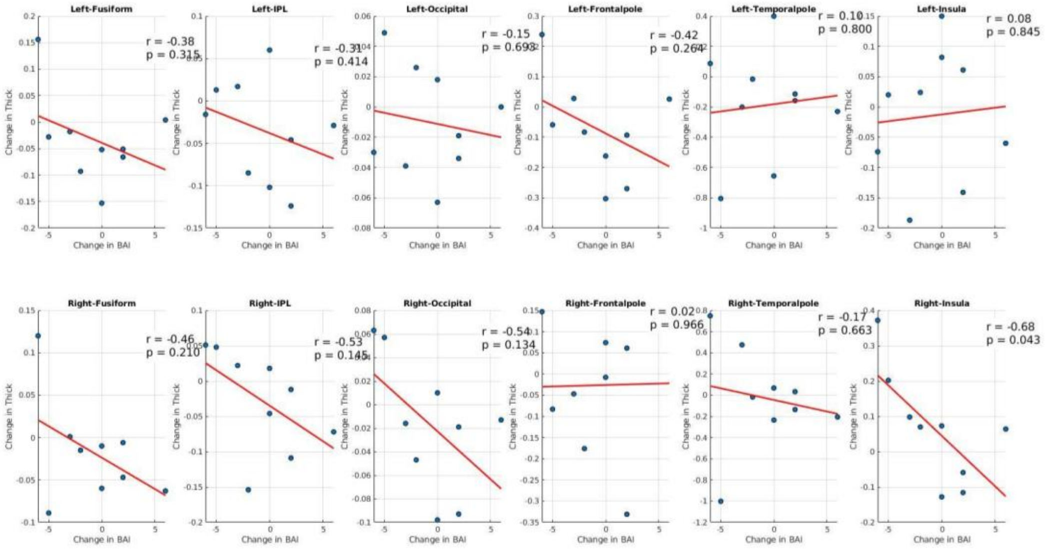
Left-Fusiform
r = -0.38
p = 0.315
Left-IPL
r = -0.31
p = 0.414
Left-Occipital
r = -0.15
p = 0.698
Left-Frontalpole
r = -0.42
p = 0.264
Left-Temporalpole
r = 0.10
p = 0.800
Left-Insula
r = 0.08
p = 0.845
Right-Fusiform
r = -0.46
p = 0.210
Right-IPL
r = -0.53
p = 0.145
Right-Occipital
r = -0.54
p = 0.134
Right-Frontalpole
r = 0.02
p = 0.966
Right-Temporalpole
r = -0.17
p = 0.663
Right-Insula
r = -0.68
p = 0.043
Change in Thick
Change in BAI

## c) FA

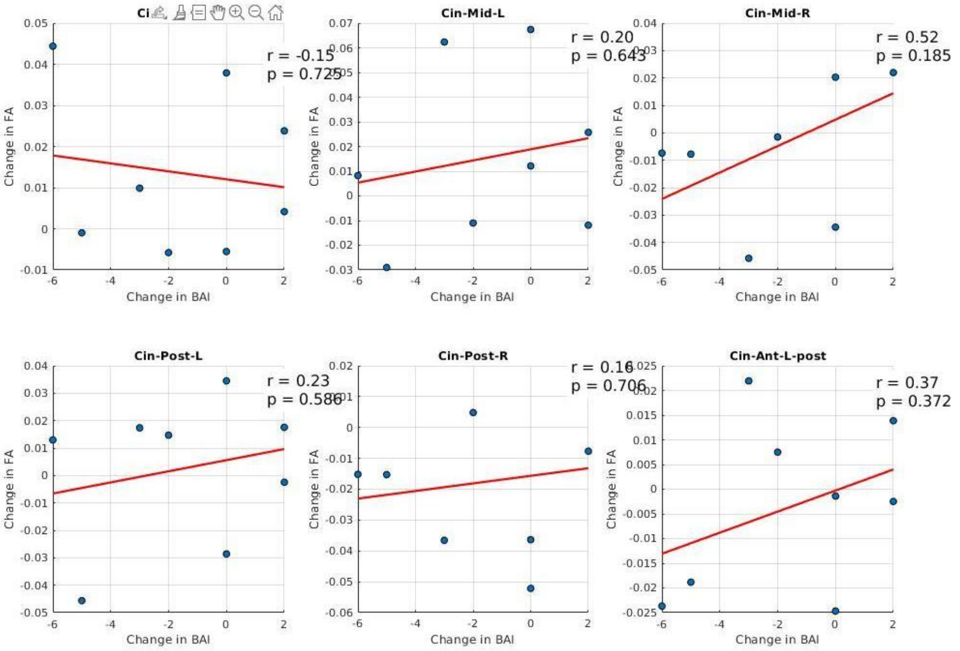
r = -0.15
p = 0.725
Cin-Mid-L
r = 0.20
p = 0.643
Cin-Mid-R
r = 0.52
p = 0.185
Cin-Post-L
r = 0.23
p = 0.586
Cin-Post-R
r = 0.16
p = 0.706
Cin-Ant-L-post
r = 0.37
p = 0.372
Change in FA
Change in BAI

### d) AAL

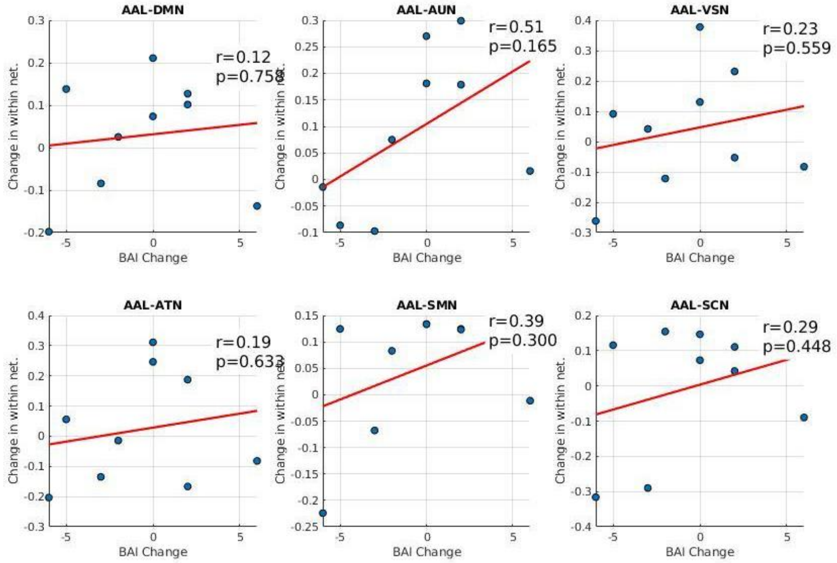


### e) Yeo

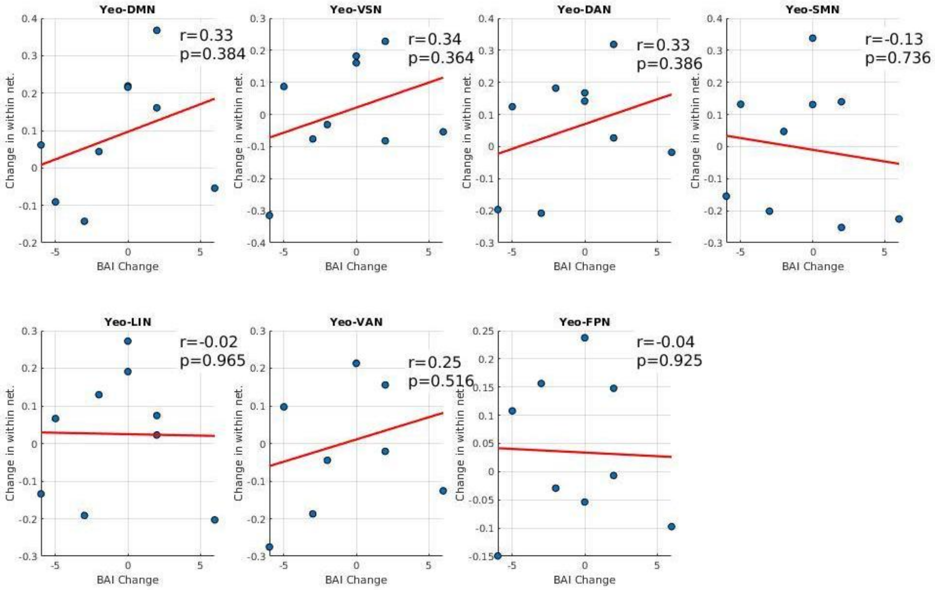


2- Severe anxiety group (BAI > 15)

### a) Subcortical volume

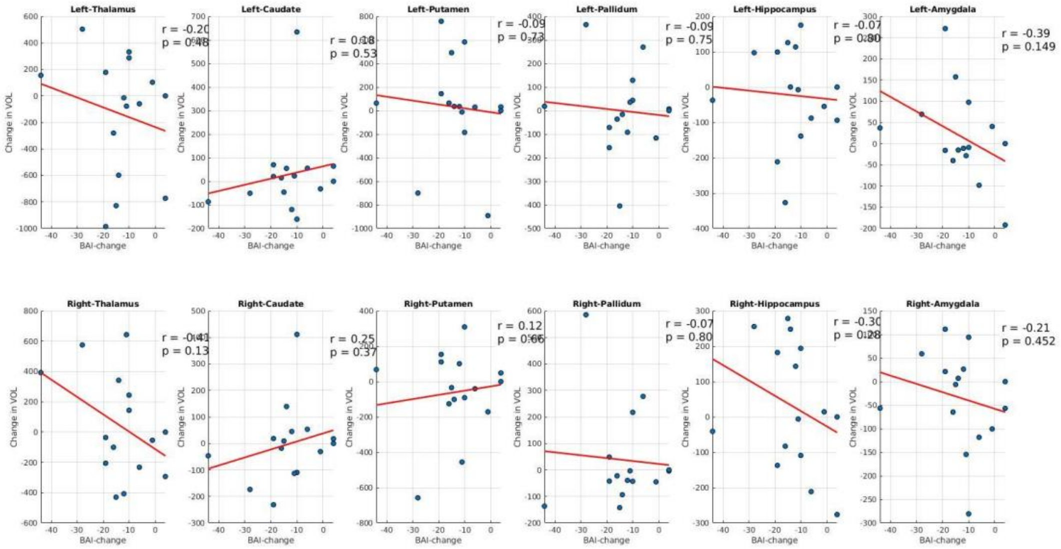


**b) Cortical Thickness**

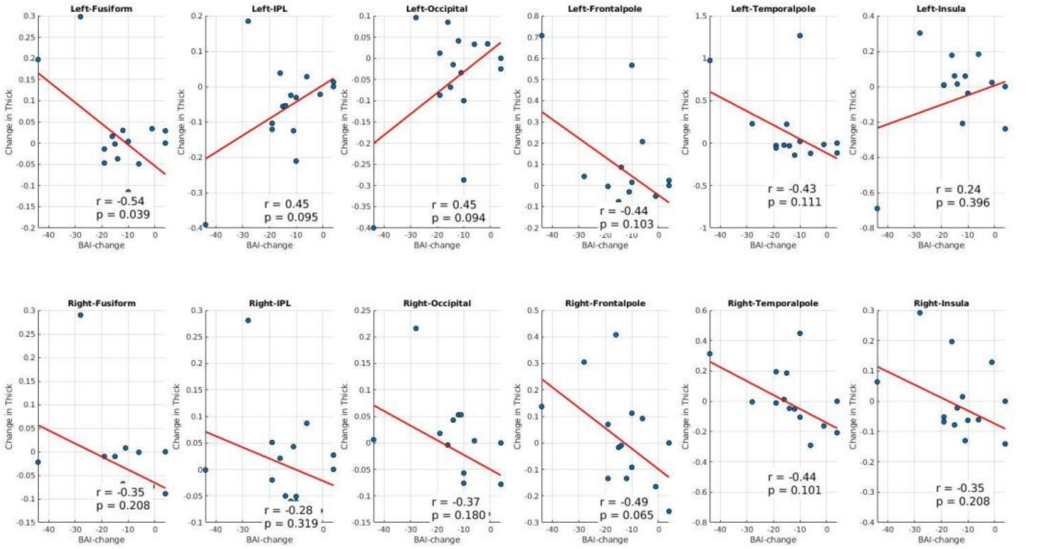


**c) FA**

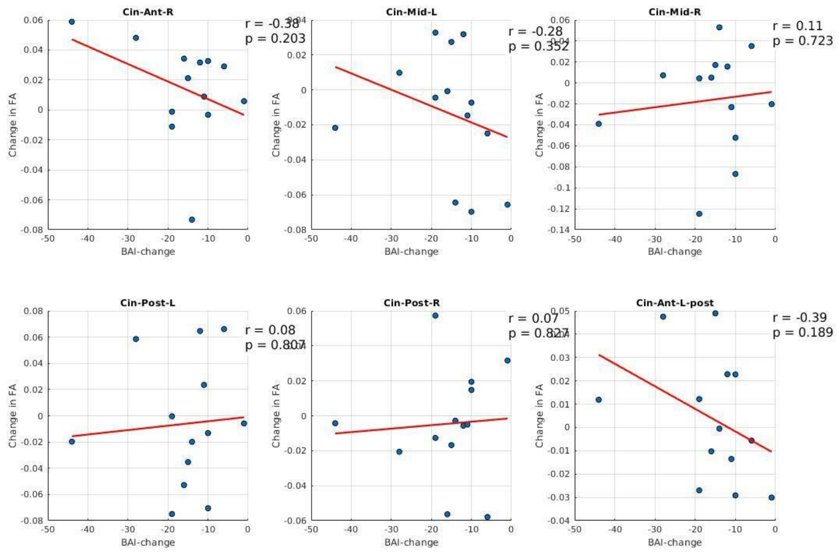
Cin-Ant-R
r = -0.38
p = 0.203
Cin-Mid-L
r = -0.28
p = 0.352
Cin-Mid-R
r = 0.11
p = 0.723
Cin-Post-L
r = 0.08
p = 0.807
Cin-Post-R
r = 0.07
p = 0.827
Cin-Ant-L-post
r = -0.39
p = 0.189
Change in FA
BAI-change

**d) AAL**

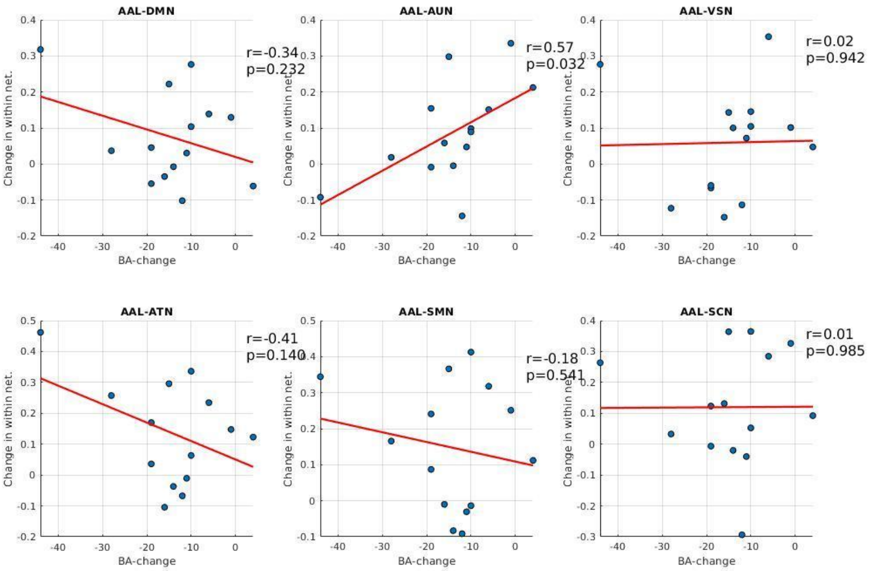
AAL-DMN
r=-0.34
p=0.232
AAL-AUN
r=0.57
p=0.032
AAL-VSN
r=0.02
p=0.942
AAL-ATN
r=-0.41
p=0.140
AAL-SMN
r=-0.18
p=0.541
AAL-SCN
r=0.01
p=0.985
Change in within net.
BA-change

**e) Yeo**

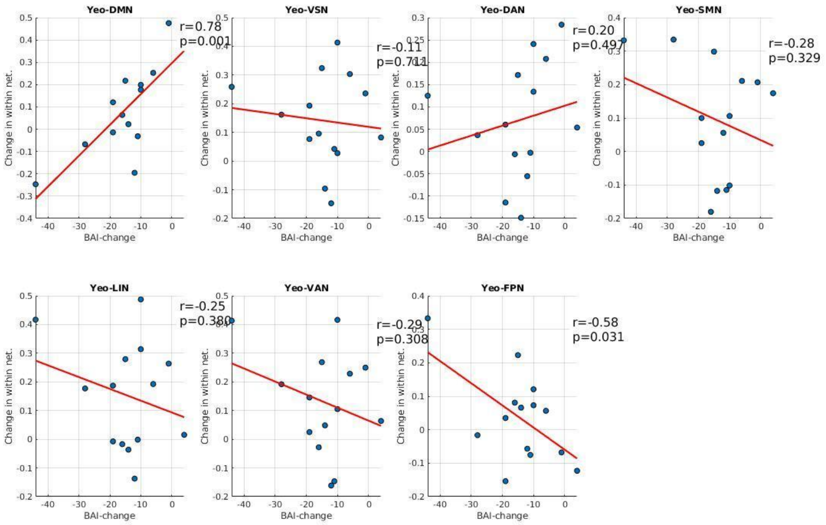


Figure S5. Association between changes in anxiety scores and changes in MRI measures. This figure displays correlation coefficients (r) between changes in anxiety scores (BAI-post - BAI-pre) and changes in structural and functional MRI indices across two patient subgroups stratified by baseline symptom severity. Negative ΔBAI values indicate improvement in anxiety symptoms. Positive and negative correlation coefficients indicate direct and inverse associations, respectively. (1) Mild anxiety group (BAI-pre < 15): Correlation coefficients between BAI changes and changes in MRI measures for patients with mild baseline anxiety symptoms: (a) Subcortical volumes; (b) Cortical thickness; (c) Fractional anisotropy (FA); (d) Within-network functional connectivity based on AAL atlas; (e) Within-network functional connectivity based on Yeo atlas. (2) Severe anxiety group (BAI-pre ≥ 15): Correlation coefficients between ΔBAI and changes in MRI measures for patients with severe baseline anxiety symptoms: (a) Subcortical volumes; (b) Cortical thickness; (c) Fractional anisotropy (FA); (d) Within-network functional connectivity based on AAL atlas; (e) Within-network functional connectivity based on Yeo atlas.

**Table S1. Brain regions and network assignments.** Column 1: index; Column 2: region name; Columns 3–5: stereotaxic (x, y, z) coordinates; Column 6: AAL network label; Column 7: AAL network ID; Column 8: Yeo network label; Column 9: Yeo network ID. *Notes:* AAL, Automated Anatomical Labeling; Yeo networks, Yeo et al. parcellation (7-network solution).

| No | Name | X | Y | Z | AAL Network | AAL | Yeo Network | Yeo |
|---|---|---|---|---|---|---|---|---|
| 1 | Precentral_L | −39 | −6 | 51 | Sensorimotor Network | 5 | Sensorimotor Network | 2 |
| 2 | Precentral_R | 41 | −8 | 52 | Sensorimotor Network | 5 | Sensorimotor Network | 2 |
| 3 | Frontal_Sup_L | −18 | 35 | 42 | Attention Network | 4 | Frontoparietal Network | 6 |
| 4 | Frontal_Sup_R | 22 | 31 | 44 | Attention Network | 4 | Frontoparietal Network | 6 |
| 5 | Frontal_Sup_Orb_L | −17 | 47 | −13 | Attention Network | 4 | Default Mode Network | 7 |
| 6 | Frontal_Sup_Orb_R | 18 | 48 | −14 | Attention Network | 4 | Default Mode Network | 7 |
| 7 | Frontal_Mid_L | −33 | 33 | 35 | Attention Network | 4 | Attention Network | 4 |
| 8 | Frontal_Mid_R | 38 | 33 | 34 | Attention Network | 4 | Attention Network | 4 |
| 9 | Frontal_Mid_Orb_L | −31 | 50 | −10 | Attention Network | 4 | Frontoparietal Network | 6 |
| 10 | Frontal_Mid_Orb_R | 33 | 53 | −11 | Attention Network | 4 | Frontoparietal Network | 6 |
| 11 | Frontal_Inf_Oper_L | −48 | 13 | 19 | Attention Network | 4 | Frontoparietal Network | 6 |
| 12 | Frontal_Inf_Oper_R | 50 | 15 | 21 | Attention Network | 4 | Frontoparietal Network | 6 |
| 13 | Frontal_Inf_Tri_L | −46 | 30 | 14 | Attention Network | 4 | Attention Network | 4 |
| 14 | Frontal_Inf_Tri_R | 50 | 30 | 14 | Attention Network | 4 | Attention Network | 4 |
| 15 | Frontal_Inf_Orb_L | −36 | 31 | −12 | Attention Network | 4 | Limbic Network | 5 |
| 16 | Frontal_Inf_Orb_R | 41 | 32 | −12 | Attention Network | 4 | Limbic Network | 5 |
| 17 | Rolandic_Oper_L | −47 | −8 | 14 | -------- | --- | Sensorimotor Network | 2 |
| 18 | Rolandic_Oper_R | 53 | −6 | 15 | -------- | --- | Sensorimotor Network | 2 |
| 19 | Supp_Motor_Area_L | −5 | 5 | 61 | Sensorimotor Network | 5 | Sensorimotor Network | 2 |
| 20 | Supp_Motor_Area_R | 9 | 0 | 62 | Sensorimotor Network | 5 | Sensorimotor Network | 2 |
| 21 | Olfactory_L | −8 | 15 | −11 | -------- | 0 | Default Mode Network | 7 |
| 22 | Olfactory_R | 10 | 16 | −11 | -------- | 0 | Default Mode Network | 7 |
| 23 | Frontal_Sup_Medial_L | −5 | 49 | 31 | Default Mode Network | 1 | Default Mode Network | 7 |
| 24 | Frontal_Sup_Medial_R | 9 | 51 | 30 | Default Mode Network | 1 | Default Mode Network | 7 |
| 25 | Frontal_Med_Orb_L | −5 | 54 | −7 | Default Mode Network | 1 | Default Mode Network | 7 |
| 26 | Frontal_Med_Orb_R | 8 | 52 | −7 | Default Mode Network | 1 | Default Mode Network | 7 |

| No | Name | X | Y | Z | AAL Network | AAL | Yeo Network | Yeo |
|---|---|---|---|---|---|---|---|---|
| 27 | Rectus_L | −5 | 37 | −18 | -------- | 0 | Default Mode Network | 7 |
| 28 | Rectus_R | 8 | 36 | −18 | -------- | 0 | Default Mode Network | 7 |
| 29 | Insula_L | −35 | 7 | 3 | Attention Network | 4 | Frontoparietal Network | 6 |
| 30 | Insula_R | 39 | 6 | 2 | Attention Network | 4 | Frontoparietal Network | 6 |
| 31 | Cingulum_Ant_L | −4 | 35 | 14 | Default Mode Network | 1 | Default Mode Network | 7 |
| 32 | Cingulum_Ant_R | 8 | 37 | 16 | Default Mode Network | 1 | Default Mode Network | 7 |
| 33 | Cingulum_Mid_L | −5 | −15 | 42 | Default Mode Network | 1 | Frontoparietal Network | 6 |
| 34 | Cingulum_Mid_R | 8 | −9 | 40 | Default Mode Network | 1 | Frontoparietal Network | 6 |
| 35 | Cingulum_Post_L | −5 | −43 | 25 | Default Mode Network | 1 | Default Mode Network | 7 |
| 36 | Cingulum_Post_R | 7 | −42 | 22 | Default Mode Network | 1 | Default Mode Network | 7 |
| 37 | Hippocampus_L | −25 | −21 | −10 | Default Mode Network | 1 | Dorsal Attention Network | 3 |
| 38 | Hippocampus_R | 29 | −20 | −10 | Default Mode Network | 1 | Dorsal Attention Network | 3 |
| 39 | ParaHippocampal_L | −21 | −16 | −21 | Default Mode Network | 1 | Limbic Network | 5 |
| 40 | ParaHippocampal_R | 25 | −15 | −20 | Default Mode Network | 1 | Limbic Network | 5 |
| 41 | Amygdala_L | −23 | −1 | −17 | -------- | 0 | Dorsal Attention Network | 3 |
| 42 | Amygdala_R | 27 | 1 | −18 | -------- | 0 | Dorsal Attention Network | 3 |
| 43 | Calcarine_L | −7 | −79 | 6 | Visual Network | 2 | Visual Attention Network | 1 |
| 44 | Calcarine_R | 16 | −73 | 9 | Visual Network | 2 | Visual Attention Network | 1 |
| 45 | Cuneus_L | −6 | −80 | 27 | -------- | 0 | Visual Attention Network | 1 |
| 46 | Cuneus_R | 14 | −79 | 28 | -------- | 0 | Visual Attention Network | 1 |
| 47 | Lingual_L | −15 | −68 | −5 | Visual Network | 2 | Visual Attention Network | 1 |
| 48 | Lingual_R | 16 | −67 | −4 | Visual Network | 2 | Visual Attention Network | 1 |
| 49 | Occipital_Sup_L | −17 | −84 | 28 | Visual Network | 2 | Visual Attention Network | 1 |
| 50 | Occipital_Sup_R | 24 | −81 | 31 | Visual Network | 2 | Visual Attention Network | 1 |
| 51 | Occipital_Mid_L | −32 | −81 | 16 | Visual Network | 2 | Visual Attention Network | 1 |
| 52 | Occipital_Mid_R | 37 | −80 | 19 | Visual Network | 2 | Visual Attention Network | 1 |
| 53 | Occipital_Inf_L | −36 | −78 | −8 | Visual Network | 2 | Visual Attention Network | 1 |
| 54 | Occipital_Inf_R | 38 | −82 | −8 | Visual Network | 2 | Visual Attention Network | 1 |
| 55 | Fusiform_L | −31 | −40 | −20 | Visual Network | 2 | Limbic Network | 5 |
| 56 | Fusiform_R | 34 | −39 | −20 | Visual Network | 2 | Limbic Network | 5 |
| 57 | Postcentral_L | −42 | −23 | 49 | Sensorimotor Network | 5 | Sensorimotor Network | 2 |

| No | Name | X | Y | Z | AAL Network | AAL | Yeo Network | Yeo |
|---|---|---|---|---|---|---|---|---|
| 58 | Postcentral_R | 41 | −25 | 53 | Sensorimotor Network | 5 | Sensorimotor Network | 2 |
| 59 | Parietal_Sup_L | −23 | −60 | 59 | Sensorimotor Network | 5 | Dorsal Attention Network | 3 |
| 60 | Parietal_Sup_R | 26 | −59 | 62 | Sensorimotor Network | 5 | Dorsal Attention Network | 3 |
| 61 | Parietal_Inf_L | −43 | −46 | 47 | Default Mode Network | 1 | Dorsal Attention Network | 3 |
| 62 | Parietal_Inf_R | 46 | −46 | 50 | Default Mode Network | 1 | Dorsal Attention Network | 3 |
| 63 | SupraMarginal_L | −56 | −34 | 30 | Sensorimotor Network | 5 | Sensorimotor Network | 2 |
| 64 | SupraMarginal_R | 58 | −32 | 34 | Sensorimotor Network | 5 | Sensorimotor Network | 2 |
| 65 | Angular_L | −44 | −61 | 36 | Default Mode Network | 1 | Default Mode Network | 7 |
| 66 | Angular_R | 46 | −60 | 39 | Default Mode Network | 1 | Default Mode Network | 7 |
| 67 | Precuneus_L | −7 | −56 | 48 | Visual Network | 2 | Dorsal Attention Network | 3 |
| 68 | Precuneus_R | 10 | −56 | 44 | Visual Network | 2 | Dorsal Attention Network | 3 |
| 69 | Paracentral_Lobule_L | −8 | −25 | 70 | Sensorimotor Network | 5 | Sensorimotor Network | 2 |
| 70 | Paracentral_Lobule_R | 7 | −32 | 68 | Sensorimotor Network | 5 | Sensorimotor Network | 2 |
| 71 | Caudate_L | −11 | 11 | 9 | Subcortical Network | 3 | Subcortical Network | 0 |
| 72 | Caudate_R | 15 | 12 | 9 | Subcortical Network | 3 | Subcortical Network | 0 |
| 73 | Putamen_L | −24 | 4 | 2 | Subcortical Network | 3 | Subcortical Network | 0 |
| 74 | Putamen_R | 28 | 5 | 2 | Subcortical Network | 3 | Subcortical Network | 0 |
| 75 | Pallidum_L | −18 | 0 | 0 | Subcortical Network | 3 | Subcortical Network | 0 |
| 76 | Pallidum_R | 21 | 0 | 0 | Subcortical Network | 3 | Subcortical Network | 0 |
| 77 | Thalamus_L | −11 | −18 | 8 | Subcortical Network | 3 | Subcortical Network | 0 |
| 78 | Thalamus_R | 13 | −18 | 8 | Subcortical Network | 3 | Subcortical Network | 0 |
| 79 | Heschl_L | −42 | −19 | 10 | -------- | 0 | Frontoparietal Network | 6 |
| 80 | Heschl_R | 46 | −17 | 10 | -------- | 0 | Frontoparietal Network | 6 |
| 81 | Temporal_Sup_L | −53 | −21 | 7 | Auditory Network | 6 | Frontoparietal Network | 6 |
| 82 | Temporal_Sup_R | 58 | −22 | 7 | Auditory Network | 6 | Frontoparietal Network | 6 |
| 83 | Temporal_Pole_Sup_L | −40 | 15 | −20 | Auditory Network | 6 | Limbic Network | 5 |
| 84 | Temporal_Pole_Sup_R | 48 | 15 | −17 | Auditory Network | 6 | Limbic Network | 5 |
| 85 | Temporal_Mid_L | −56 | −34 | −2 | Default Mode Network | 1 | Frontoparietal Network | 6 |
| 86 | Temporal_Mid_R | 57 | −37 | −1 | Default Mode Network | 1 | Frontoparietal Network | 6 |
| 87 | Temporal_Pole_Mid_L | −36 | 15 | −34 | Default Mode Network | 1 | Default Mode Network | 7 |
| 88 | Temporal_Pole_Mid_R | 44 | 15 | −32 | Default Mode Network | 1 | Default Mode Network | 7 |

| No | Name | X | Y | Z | AAL Network | AAL | Yeo Network | Yeo |
|---|---|---|---|---|---|---|---|---|
| 89 | Temporal_Inf_L | −50 | −28 | −23 | Attention Network | 4 | Default Mode Network | 7 |
| 90 | Temporal_Inf_R | 54 | −31 | −22 | Attention Network | 4 | Default Mode Network | 7 |

Table S2. The estimated Repeatability Coefficient of subcortical volume, cortical thickness, FA, and within-network connectivity. RC, repeatability coefficient; $RC_L$, Lower RC; $RC_U$, Upper RC; IPL, inferior parietal lobule.

| ***Subcortical region*** | ***$RC_L$*** | ***$RC_U$*** | ***RC*** |
|---|---|---|---|
| *Left-Thalamus* | 0.0768 | 0.1449 | 0.1003 |
| *Left-Caudate* | 0.0718 | 0.1354 | 0.0938 |
| *Left-Putamen* | 0.0890 | 0.1679 | 0.1163 |
| *Left-Pallidum* | 0.0794 | 0.1499 | 0.1038 |
| *Left-Hippocampus* | 0.0865 | 0.1632 | 0.1130 |
| *Left-Amygdala* | 0.1339 | 0.2527 | 0.1750 |
| *Right-Thalamus* | 0.0669 | 0.1262 | 0.0874 |
| *Right-Caudate* | 0.0614 | 0.1160 | 0.0803 |
| *Right-Putamen* | 0.0700 | 0.1322 | 0.0915 |
| *Right-Pallidum* | 0.0890 | 0.1680 | 0.1164 |
| *Right-Hippocampus* | 0.1086 | 0.2050 | 0.1420 |
| *Right-Amygdala* | 0.1269 | 0.2394 | 0.1658 |
| ***Cortical region*** | ***$RC_L$*** | ***$RC_U$*** | ***RC*** |
| *Left-Fusiform* | 0.0401 | 0.0757 | 0.0524 |
| *Left-IPL* | 0.0288 | 0.0544 | 0.0377 |
| *Left-Occipital* | 0.0270 | 0.0509 | 0.0353 |
| *Left-Frontalpole* | 0.0769 | 0.1451 | 0.1005 |
| *Left-Temporalpole* | 0.1285 | 0.2426 | 0.1680 |
| *Left-Insula* | 0.0571 | 0.1079 | 0.0747 |
| *Right-Fusiform* | 0.0469 | 0.0885 | 0.0613 |
| *Right-IPL* | 0.0372 | 0.0702 | 0.0486 |
| *Right-Occiptal* | 0.0334 | 0.0630 | 0.0436 |
| *Right-Frontalpole* | 0.066 | 0.1247 | 0.0863 |
| *Right-Temporalpole* | 0.119 | 0.2246 | 0.1555 |
| *Right-Insula* | 0.0381 | 0.0719 | 0.0498 |
| ***White matter regions*** | ***$RC_L$*** | ***$RC_U$*** | ***RC*** |
| *Cingulum-Ant-L* | 0.2075 | 0.3986 | 0.2729 |

| | | | |
|---|---|---|---|
| *Cingulum-Ant-R* | 0.4787 | 0.9194 | 0.6295 |
| *Cingulum-Mid-L* | 0.0859 | 0.1649 | 0.1129 |
| *Cingulum-Mid-R* | 0.0941 | 0.1808 | 0.1238 |
| *Cingulum-Post-L* | 0.1253 | 0.2407 | 0.1648 |
| *Cingulum-Post-R* | 0.1006 | 0.1933 | 0.1323 |
| ***AAL networks*** | ***$RC_L$*** | ***$RC_U$*** | ***RC*** |
| *Default-mode* | 0.1570 | 0.2963 | 0.2052 |
| *Auditory* | 0.1344 | 0.2537 | 0.1757 |
| *Visual* | 0.0955 | 0.1803 | 0.1249 |
| *Attention* | 0.1820 | 0.3434 | 0.2378 |
| *Sensorimotor* | 0.2075 | 0.3917 | 0.2713 |
| *Subcortical* | 0.1063 | 0.2006 | 0.1389 |
| ***Yeo networks*** | ***$RC_L$*** | ***$RC_U$*** | ***RC*** |
| *Default-mode* | 0.1594 | 0.3009 | 0.2083 |
| *Visual* | 0.2252 | 0.4251 | 0.2944 |
| *Dorsal attention* | 0.1252 | 0.2362 | 0.1636 |
| *Sensorimotor* | 0.0739 | 0.1396 | 0.0967 |
| *Limbic* | 0.1263 | 0.2384 | 0.1651 |
| *Ventral attention* | 0.2114 | 0.3990 | 0.2763 |
| *Fronto-parietal* | 0.1961 | 0.3702 | 0.2564 |
